# Life after mild traumatic brain injury: Widespread structural brain changes associated with psychological distress revealed with multimodal magnetic resonance imaging

**DOI:** 10.1101/2021.11.03.21265823

**Authors:** Francesca Sibilia, Rachel M. Custer, Andrei Irimia, Farshid Sepehrband, Arthur Toga, Ryan P. Cabeen, the TRACK-TBI Investigators

## Abstract

**Background:** Traumatic brain injury (TBI) can alter brain structure and lead to onset of persistent neuropsychological symptoms. This study investigates the relationship between brain damage and psychological distress after mild TBI (mTBI) using multimodal magnetic resonance imaging (MRI).

**Methods:** Eighty-nine mTBI patients from the TRACK-TBI (Transforming Research and Clinical Knowledge in Traumatic Brain Injury) pilot study were included. Subscales of the Brief Symptoms Inventory 18 for depression, anxiety, and somatization were used as outcome measures of psychological distress ∼6 months after the traumatic event. Glasgow Coma Scale scores were used to evaluate recovery. MRIs were acquired within 2 weeks post-injury. Perivascular spaces (PVS) were segmented using an enhanced PVS segmentation method, and the volume fraction was calculated for the whole brain and white matter regions. Cortical thickness and gray matter structures volumes were calculated in Freesurfer; diffusion imaging indices and multi-fiber tracts were extracted using the Quantitative Imaging Toolkit. The analysis was performed considering age, sex, intracranial volume, educational attainment, and improvement level upon discharge as covariates.

**Results:** Perivascular space fractions in the posterior cingulate, fusiform, and postcentral areas were found to be associated with somatization symptoms. Depression, anxiety, and somatization symptoms were associated with the cortical thickness of the frontal- opercularis and occipital pole, putamen and amygdala volumes, and corticospinal tract and superior thalamic radiation. Analyses were also performed on the two hemispheres separately to explore lateralization.

**Conclusions:** This study shows how PVS, cortical, and microstructural damages can predict the onset of depression, anxiety, and somatization symptoms in mTBI patients.

## 1. Introduction

Traumatic brain injury (TBI) is defined as a nondegenerative, noncongenital insult to the brain from an external mechanical force. Based on the duration of loss of consciousness and post-traumatic amnesia, TBI can be defined as mild, moderate, and severe, measured via the Glasgow Coma Scale (GCS) [1]. A GCS score between 13-15 defines mild TBI (mTBI), GCS scores from 9 to 12 indicate moderate TBI, and scores of GCS ≤ 8 indicate severe TBI [2]. In mTBI, which represents 70% of all TBI cases [3], loss of consciousness is ∼30 minutes or less, and posttraumatic amnesia (PTA) is less than 24 hours [4], leading to potential cognitive, physical, and psychological impairments. There is evidence of cognitive and physical post-injury improvement [5] [6], while less is known about the psychological consequences in mTBI sub-acute stages (>3 months). Psychological distress is defined as a “unique discomforting, emotional state experienced by an individual in response to a specific stressor or demand that results in harm, either temporary or permanent, to the person” [7].

The onset of psychological symptoms in mTBI is usually seen 24-72 hours after the traumatic event, with improvements within 3 months from injury [8]. The risk of developing depression and anxiety can persist after acute stages [9][10][11]. The onset of psychological symptoms is therefore an important predictor for monitoring recovery within 6 months following TBI [12]. The emotional dysregulation following TBI reflects a potential neural injury that can be detected with neuroimaging techniques, such as magnetic resonance imaging (MRI) [12]. In recent years, automated neuroimaging analysis tools have been used to study structural damages in cortical thickness, white matter (WM) microstructure, and vascular components following mTBI [13]. Most studies focused on a single type of structural alteration, but it is likely that the onset of psychological distress is caused by simultaneous damage across multiple brain components. Addressing this issue requires further investigation of multiple distinct brain systems in the same cohort with respect to a wider range of measures of psychological stress and well-being.

The aim of this study is to identify anatomical associations with depressive, anxiety, and somatization symptoms in mTBI patients six months after the traumatic event. We used a multimodal structural neuroimaging approach that provides a combined view of cortical morphometry, WM connectivity, and perivascular spaces. We further investigated the relationship between the structural MRI measures and the level of life satisfaction within the first 6 months post-TBI. The onset of psychological distress can be influenced by the level of happiness perceived, as shown by previous studies [14][15]; nevertheless, the association between structural imaging parameters and life satisfaction in TBI patients is still unclear. Investigating such a relationship is crucial to understanding the underlying neural disruptions that can affect quality of life in mTBI patients.

## 2. Methods and Materials

### 2.1. Participants

Participants (n=89, age range: 16-80 years old) were part of the Transforming Research and Clinical Knowledge in Traumatic Brain Injury (TRACK-TBI) pilot study. TRACK-TBI is a prospective, multicenter observational study conducted at 18 U.S. trauma centers, enrolling patients with TBI between February 26, 2014, and August 8, 2018. Demographic, psychological assessment, and imaging data were downloaded from the Federal Interagency Traumatic Brain Injury Research (FITBIR) repository (https://fitbir.nih.gov).

Inclusion criteria for the TRACK-TBI Pilot were an age over 16 years old, acute external force trauma to the head, presentation to an enrolling center, and performance of non-contrast head CT to assess for evidence of acute TBI within 24 hours of injury. Exclusion criteria were pregnancy, ongoing life-threatening disease (e.g., end-stage malignancy), police custody, involuntary psychiatric hold, and non-English speakers (as multiple outcome measures were administered and/or normed only in English). Participants were mostly White, and right-handed. All study protocols were approved by the University of California at San Francisco Institutional Review Board, and all patients and control subjects or their legal representatives gave written informed consent.

### 2.2. Cognitive assessment

Demographic information, clinical measures, and scores of psychological distress subgroups were collected for each participant. Glasgow Coma Scale is used to rate consciousness levels following TBI. The total score is the result of three components (verbal, motor, and eye responses), which determine the severity of the traumatic event. It is usually used in the field upon emergency department (ED) arrival and at the time of discharge [16]. In this study, GCS scores between 13-15 were used to determine the level of improvement for each participant, calculated via the difference in discharge and ED GCS scores.

#### 2.2.1. Brief Symptoms Inventory 18 (BSI-18)

The Brief Symptom Inventory 18 (BSI-18) quantifies psychological distress and comorbidities in patients with mental and somatic illnesses [17]. The BSI-18 is the shortened version of the BSI [18]. It is reduced to three subscales (Anxiety, Depression and Somatization) formed by six items each and the Global Severity Index (GSI), to improve dimensionality structure and be more accessible to patients [19]. Scores for each item are summed up across participants to obtain the total scores for Anxiety, Depression, and Somatization, and then converted to T-scores. The item list for each subscale is found in Table S1. The GSI total score ranges between 0 – 72 and the three scales scores range between 0 – 24. The BSI-18 manual suggests respondents with a total GSI t-score ≥ 63 as having significant psychological distress [20].

#### 2.2.2. Life satisfaction measurement (SWLS)

Life satisfaction is defined as “a global assessment of a person’s quality of life according to his chosen criteria” [21], based on a cognitive and judgmental process rather than emotional evaluation [22]. The Satisfaction with Life scale (SWLS) is a 5-item questionnaire where participants are asked to rate the level of satisfaction and conditions of their lives as whole. Outcomes are categorized as ‘slightly dissatisfied/satisfied’, ‘dissatisfied/satisfied’, ‘extremely dissatisfied/satisfied’. A score of 20 represents the neutral point on the scale [23].

### 2.3. MRI data acquisition

3T MRI sequences were obtained in a SIGNA EXCITE scanner with 8-channel coil. Axial three-dimensional (3D) inversion recovery fast spoiled gradient recalled echo T1- weighted images were acquired with TE = 1.5 ms; TR = 6.3 ms; inversion time = 400 ms; flip angle, 15 degrees, with 230-mm FOV, 156 contiguous partitions of 1 mm and matrix size 256x256 [24].

Diffusion images (DWI) were acquired with a multi-slice single-shot spin echo echo-planar pulse sequence (echo time [TE] = 63 ms; repetition time [TR] = 14 sec) using 55 diffusion-encoding directions, acquired at b = 1000 sec/mm2, seven acquisitions at b = 0 sec/mm2, 72 inter- leaved slices of 1.8-mm thickness and no gap between slices, a 128x128 matrix, and a field of view (FOV) of 230x230 mm.

We implemented a multi-modal image analysis pipeline that included components for white matter analysis of fiber bundles, gray matter analysis of cortical thickness and subcortical volume, and perivascular space analysis to detect abnormal PVS enlargement in the brain vasculature. The steps of the pipeline are described as follows.

### 2.4. Diffusion MRI Preprocessing

Standard DWI processing was performed using a combination of the FSL Diffusion Toolbox [25] (Jenkinson et al. 2012), ANTs (https://stnava.github.io/ANTs/) [26], and the Quantitative Imaging Toolkit (QIT) (http://cabeen.io/qitwiki; [27]). We first applied a non-local means filter to the DWI data using the VolumeFilterNLM module in QIT [28], and then *FSL-eddy* was used to correct for eddy current-induced distortions [29]. An automated quality control step was performed to quantify the DWI data quality using *FSL- eddy_quad* [30] at a subject level. Brain segmentation was performed using FSL BET [31] (Smith, 2002), and diffusion tensor imaging (DTI) parameters were estimated using weighted-linear least squares estimation [32] implemented in the VolumeTensorFit module of QIT. Parameter maps for DTI parameters were extracted and retained for quantitative analysis [33], including fractional anisotropy (FA), mean diffusivity (MD), radial diffusivity (RD), and axial diffusivity (AD). For tractography analysis, ball-and-sticks modeling was performed using the Monte Carlo Markov Chain approach in FSL BEDPOSTX [34].

### 2.5. White matter Analysis

We performed quantitative tractography analysis to characterize the microstructure of 33 fiber bundles of projection, commissural, and association connections, with separate models for the left and right hemispheres. The bundles-of-interest (BOI) included the thalamic radiations and the corpus callosum (both subdivided by cortical target), the anterior commissure, the cingulum (subdivided into dorsal and ventral parts), the fornix, the inferior longitudinal fasciculus, middle longitudinal fasciculus, the pyramidal tract, the superior longitudinal fasciculus (I, II, and III), the arcuate fasciculus, the uncinate fasciculus, and the cerebellar peduncles (middle, inferior, and superior subdivisions). We extracted fiber bundle models from each individual case using an atlas-based bundle-specific approach. We followed a similar approach to past work [35] to create a priori bundle definitions with group-averaged multi-fiber models [36] in the IIT ICBM diffusion MRI template space [37]. We manually delineated bundle seed, inclusion, exclusion, and stopping volumetric masks for each BOI based on anatomical references [38] [39]. We performed diffeomorphic registration of the subject FA maps to the IIT template FA maps using ANTs [40], and the resulting deformations were applied to transform each bundle delineation mask into subject native space. We then applied a hybrid reinforcement tractography approach [41] to obtain subject-specific fiber bundles using the ball-and-sticks voxel-wise models; we used the following parameters: a maximum turning angle of 75 degrees, a minimum volume fraction of 0.05, trilinear interpolation, a step size of 1 mm, and a minimum length of 10 mm. We measured bundle-specific DTI parameters by sampling the voxel-wise parameter values at each curve vertex and computing the average across each entire bundle to obtain DTI metrics for statistical analysis. We also summarized each fiber bundle with its morphometric properties, e.g., mean tract density, volume, mean length, and mean bundle thickness. For each fiber bundle parameter, the average of the left and right were computed; this was considered primarily, and lateralization was investigated secondarily.

### 2.6. Gray matter analysis

Cortical surface analysis was conducted using FreeSurfer version 5.3.0 [42]. Regional averages of cortical thickness were computed using the T1-weighted MRI of each subject using the Desikan-Killiany (DK) cortical atlas [43]. Subcortical volumes were computed using the *aparc* output of the Freesurfer pipeline, which included the caudate, putamen, amygdala, thalamus, and hippocampus.

### 2.7. Perivascular space analysis

Perivascular space was segmented using a previously published technique developed in our institute [44], based on an enhanced PVS contrast (EPC).

After the data acquisition and preprocessing, T1- and T2-weighted images were filtered by using non-local mean filtering. Non-local mean technique measures the image intensity similarities by taking into account the neighboring voxels in a blockwise fashion, where filtered image is ∑_xi∈Viω_*(xi,xj)u(xj).* For each voxel (*x_j_*) the weight (*ω*) is measured using the Euclidean distance between 3D patches.

The adaptive non-local mean filtering technique adds a regularization term to the above formulation to remove bias intensity of the Rician noise observed in MRI and estimate the filter bandwidth in each voxel. This allows to remove high-frequency spatial noise at a single-voxel level and preserving signal intensities of PVS voxels. EPC was obtained by dividing filtered T1w and T2w images.

Then MRI images were parcellated using the Advanced Normalization Tools (ANTs) package [26], to obtain masks of white matter and basal ganglia. Parcellated WM was used as a mask for PVS analysis. For PVS segmentation, a Frangi filter was applied using QIT [27], which extracts the likelihood of a voxel belonging to a PVS. Frangi filter estimated vesselness measures at different scales and provided the maximum likeliness. The scale was set to a large range of 0.1 to 5 voxels in order to maximize the vessel inclusion. The output of this step was a quantitative maximum likelihood map of vessels in regions of interest. The outputs across voxels comprise vesselness measured across a range of filter scales.

A threshold was applied to the vesselness map to obtain a binary mask of PVS regions. We chose a previously optimized scaled threshold of 1.5 (equal to raw threshold of 1e- 6), which was required to calculate PVS volumetric measurements and spatial distribution.

### 2.8. Statistical analysis

Statistical analysis was performed in R version 4.0.5. Multi-variable linear regressions were performed to investigate the MRI-related parameters predicting development of psychological distress six months following TBI. A DTI-based white matter FA map from the JHU atlas [45] [46] [47], gray matter subcortical areas maps, whole-brain WM volume and cortical thickness maps from FreeSurfer, and global and region-based perivascular spaces were considered in the analysis. Diffusivity measures (FA, MD, RD and AD) and geometrical tract properties (tract density, volume, length, and thickness) from tractography results were also included.

MRI parameters presenting more than 60% of missing data across participants were removed from the analysis; in our case, the anterior commissure and the first branch of the superior longitudinal fasciculus were excluded as dependent variables in the analysis. Outliers were removed using Tukey’s method, by using twice the inter-quantile range (IQR). MRI parameters were converted into z-scores by subtracting their mean and dividing by their standard deviation, to ensure regression coefficients to be reported as standardized effect sizes.

The MRI parameters we used as dependent variable were gray matter cortical thickness, total white matter, sub-cortical structures volume, and global and regional PVS fractions based on a Freesurfer atlas. Regarding WM tracts, geometrical properties of density, length, volume, and thickness were calculated for both the entire bundle and for each tract divided into head, middle and tail. Finally, averaged DTI metrics, namely FA, MD, RD and AD were computed. BSI outcome scores for Anxiety, Depression and Somatization were used as independent variables, controlled for age, biological sex, educational attainment (in years), total intracranial volume, and improvement levels. Sex was converted into a categorical variable. Covariate missing values treatment was based on mean imputation procedure.

Uncorrected significant p-values underwent correction for multiple comparison, using Benjamini and Hochberg (BH) false discovery rate (FDR) approach [48], i.e., to obtain q-values. Adjusted R^2^ and *β* regression coefficient values were also computed and reported in the Supplementary Material.

## 3. Results

Demographic summaries are shown in Table 1. The population was formed by 60 males and 29 females, with a mean age of 37.03 ± 15.06 years old. Demographic information based on gender are shown in Table 2. In this section, results are reported considering both hemispheres together. To test lateralization effect, analyses were also run on separated hemispheres (see Supplementary Material for results).

**Table 1:** Demographic information and summary statistics of psychological distress variables and TBI-related scores in the population considered in this study. Abbreviations: Gcs = Glasgow Coma scale; ed = emergency department; Depres_T = Depression (T-score); Anx_T = Anxiety (T-score); GSI_T = General Severity Index (T-score); Somat_T = Somatization (T-score); SWLS_TOTsc = Satisfaction-with-life-scale total score.

|  | N | Mean | SD | Min | Q1 | Median | Q3 | Max |
| --- | --- | --- | --- | --- | --- | --- | --- | --- |
| age | 89 | 37.03 | 15.06 | 16.00 | 25.00 | 34.00 | 50.00 | 80.00 |
| field_gcs | 89 | 14.26 | 1.68 | 4.00 | 14.00 | 15.00 | 15.00 | 15.00 |
| discharge_gcs | 89 | 14.89 | 0.38 | 13.00 | 15.00 | 15.00 | 15.00 | 15.00 |
| ed_gcs | 89 | 14.62 | 0.67 | 11.00 | 14.00 | 15.00 | 15.00 | 15.00 |
| rivermead | 89 | 15.78 | 15.54 | 0.00 | 2.00 | 12.00 | 26.00 | 59.00 |
| EducationYear | 89 | 14.67 | 2.71 | 6.00 | 13.00 | 14.67 | 16.00 | 22.00 |
| Depres_T | 89 | 56.52 | 11.81 | 40.00 | 45.00 | 58.00 | 66.00 | 81.00 |
| Anx_T | 89 | 56.22 | 12.19 | 38.00 | 47.00 | 59.00 | 67.00 | 81.00 |
| GSI_T | 89 | 57.45 | 12.09 | 33.00 | 48.00 | 59.00 | 67.00 | 81.00 |
| Somat_T | 89 | 55.97 | 11.40 | 41.00 | 48.00 | 56.00 | 64.00 | 81.00 |
| SWLS_TOTsc | 89 | 19.59 | 7.86 | 5.00 | 14.00 | 20.00 | 25.00 | 35.00 |

**Table 2:** Demographic information of the population based on biological sex, after dummy coding (1=Male and 2= Female). Abbreviations: Gcs = Glasgow Coma scale; ed = emergency department; Depres_T = Depression (T-score); Anx_T = Anxiety (T-score); GSI_T = General Severity Index (T-score); Somat_T = Somatization (T-score); SWLS_TOTsc = Satisfaction-with-life-scale total score.

|  | sex | N | Mean | SD | Min | Q1 | Median | Q3 | Max |
| --- | --- | --- | --- | --- | --- | --- | --- | --- | --- |
| age | 1 | 60 | 38.10 | 14.73 | 16.00 | 25.00 | 37.00 | 50.00 | 80.00 |
|  | 2 | 29 | 34.83 | 15.76 | 16.00 | 23.00 | 31.00 | 48.00 | 73.00 |
| field_gcs | 1 | 60 | 14.10 | 1.96 | 4.00 | 14.00 | 15.00 | 15.00 | 15.00 |
|  | 2 | 29 | 14.59 | 0.73 | 12.00 | 14.00 | 15.00 | 15.00 | 15.00 |
| discharge_gcs | 1 | 60 | 14.92 | 0.33 | 13.00 | 15.00 | 15.00 | 15.00 | 15.00 |
|  | 2 | 29 | 14.83 | 0.47 | 13.00 | 15.00 | 15.00 | 15.00 | 15.00 |
| ed_gcs | 1 | 60 | 14.57 | 0.70 | 11.00 | 14.00 | 15.00 | 15.00 | 15.00 |
|  | 2 | 29 | 14.72 | 0.59 | 13.00 | 15.00 | 15.00 | 15.00 | 15.00 |
| rivermead | 1 | 60 | 16.18 | 15.71 | 0.00 | 2.00 | 12.50 | 26.50 | 58.00 |
|  | 2 | 29 | 14.97 | 15.44 | 0.00 | 2.00 | 9.00 | 25.00 | 59.00 |
| EducationYear | 1 | 60 | 14.70 | 2.65 | 8.00 | 13.00 | 14.67 | 16.00 | 22.00 |
|  | 2 | 29 | 14.61 | 2.87 | 6.00 | 13.00 | 15.00 | 16.00 | 20.00 |
| Depres_T | 1 | 60 | 58.52 | 12.10 | 42.00 | 48.00 | 60.00 | 67.00 | 81.00 |
|  | 2 | 29 | 52.38 | 10.16 | 40.00 | 45.00 | 50.00 | 62.00 | 70.00 |
| Anx_T | 1 | 60 | 56.72 | 12.55 | 39.00 | 47.50 | 60.00 | 68.00 | 81.00 |
|  | 2 | 29 | 55.21 | 11.57 | 38.00 | 45.00 | 57.00 | 65.00 | 74.00 |
| GSI_T | 1 | 60 | 59.10 | 11.79 | 36.00 | 50.00 | 61.00 | 68.50 | 81.00 |
|  | 2 | 29 | 54.03 | 12.20 | 33.00 | 45.00 | 57.00 | 61.00 | 79.00 |
| Somat_T | 1 | 60 | 56.88 | 11.53 | 41.00 | 48.00 | 56.00 | 65.00 | 81.00 |
|  | 2 | 29 | 54.07 | 11.10 | 41.00 | 42.00 | 50.00 | 63.00 | 77.00 |
| SWLS_TOTsc | 1 | 60 | 19.28 | 8.18 | 5.00 | 12.00 | 21.50 | 25.00 | 33.00 |
|  | 2 | 29 | 20.23 | 7.23 | 5.00 | 18.00 | 19.00 | 26.00 | 35.00 |

### 3.1. BSI-18

#### 3.1.1. Anxiety

After FDR correction, a significant negative relationship was seen between anxiety levels across the follow-up period and the cortical thickness of the inferior-opercular frontal gyrus and occipital pole, and the putamen subcortically. A significant positive relationship was seen with tract thickness of the anterior thalamic radiation (Figure 1, shown in blue), as well as with the FA of cerebral peduncle, posterior limb of internal capsule and external capsule and superior longitudinal fasciculus (SLF) from the JHU white-matter atlas (ROIs are indicated in Figure 2). Table S2 lists the brain regions significantly associated with anxiety symptoms, and Figure S1 shows the correspondent scatterplots.

**Figure 1:**
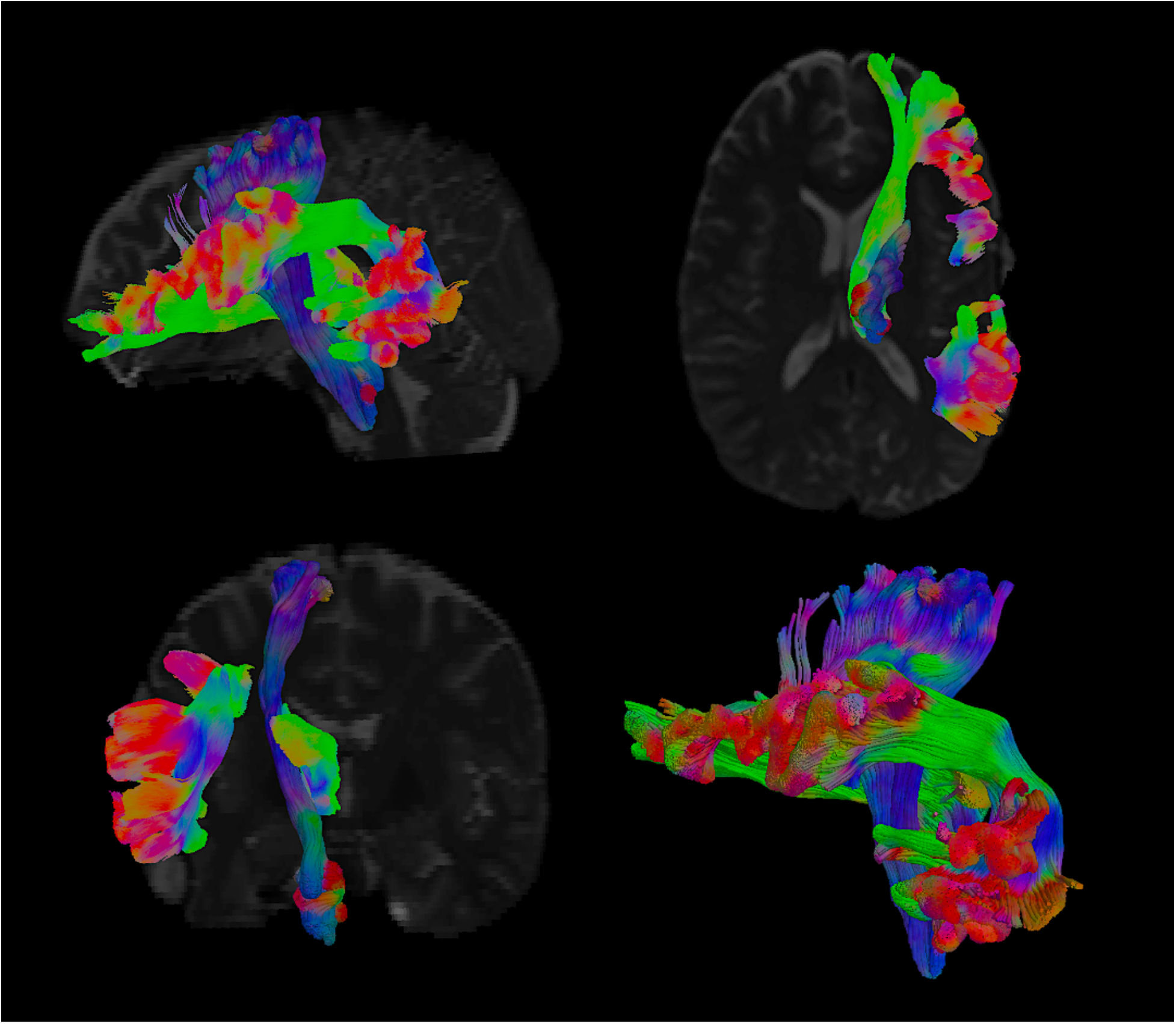
graphical representation of white matter tracts that showed geometrical features (thickness and length) significantly associated to somatization symptoms. The tract in blue (the anterior thalamic radiation) was found to be associated to both somatization and anxiety traits.

**Figure 2:**
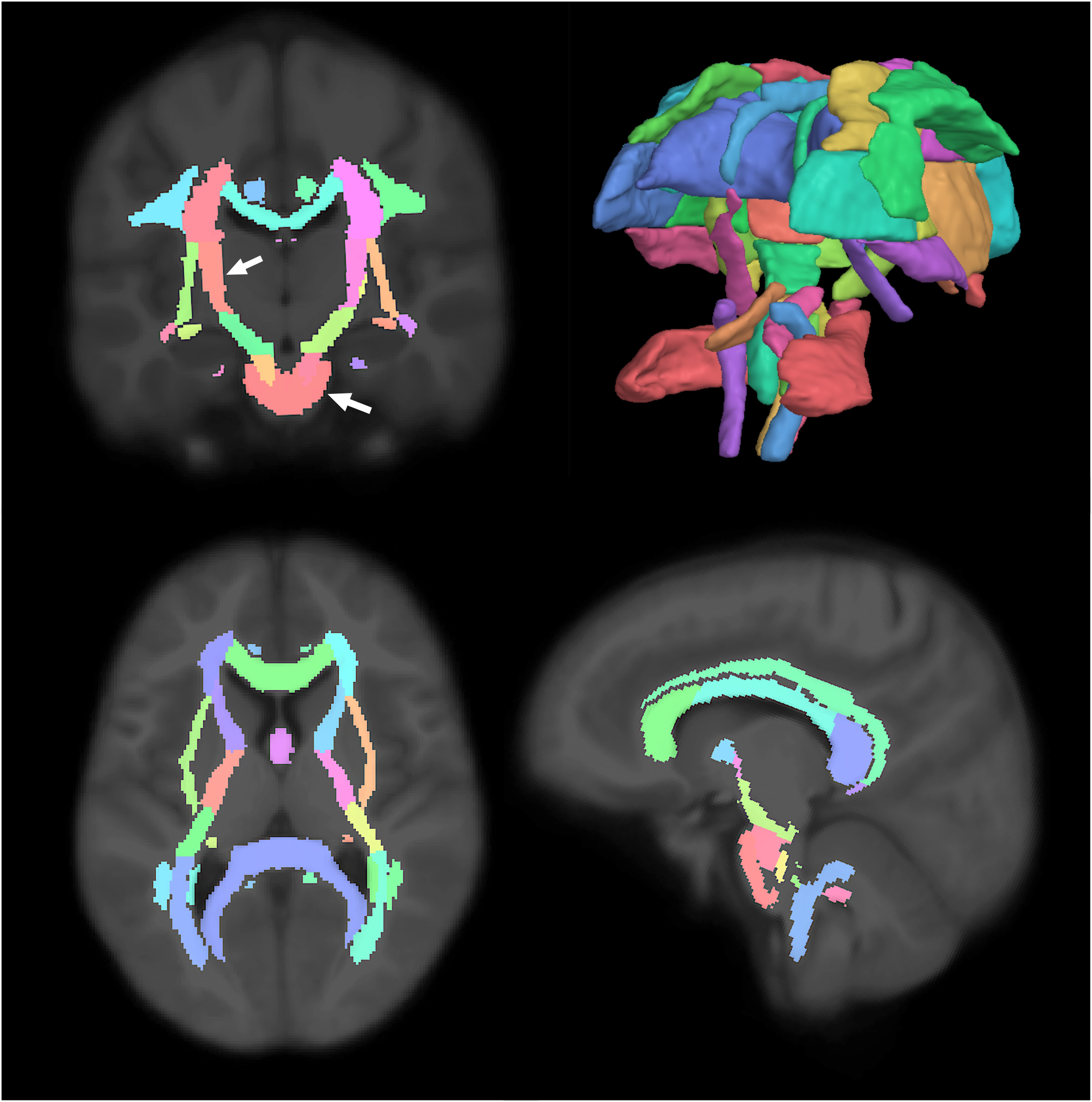
John Hopkins atlas of white matter regions. The cerebellar peduncle and internal capsule (both in “salmon” color, see arrows on the coronal view) were the common regions that showed a significant relationship between FA and the three domains of psychological distress (depression, anxiety and somatization, p<0.05 FDR corrected).

#### 3.1.2. Depression

Depression was negatively associated to widely distributed cortical thickness changes, specifically in the subcentral, frontopolar sulci and gyri, dorsal posterior cingulate, cuneus, inferior frontal opercular, fronto-superior, insular, inferior supramarginal and superior parietal gyrus, precental gyrus, middle temporal gyrus, occipital pole, circular insular sulci, medial orbito-olfactory sulcus, superior temporal sulcus (Figure 3). Fractional anisotropy was a significant predictor of psychological distress for the cerebral peduncle (*q*=0.03) and posterior limb of the internal capsule (*q*<0.001). The complete list of the significant predictors can be found in Table S3 with the respective FDR-corrected *p*-values and Figure S2 shows the correspondent scatterplots.

**Figure 3:**
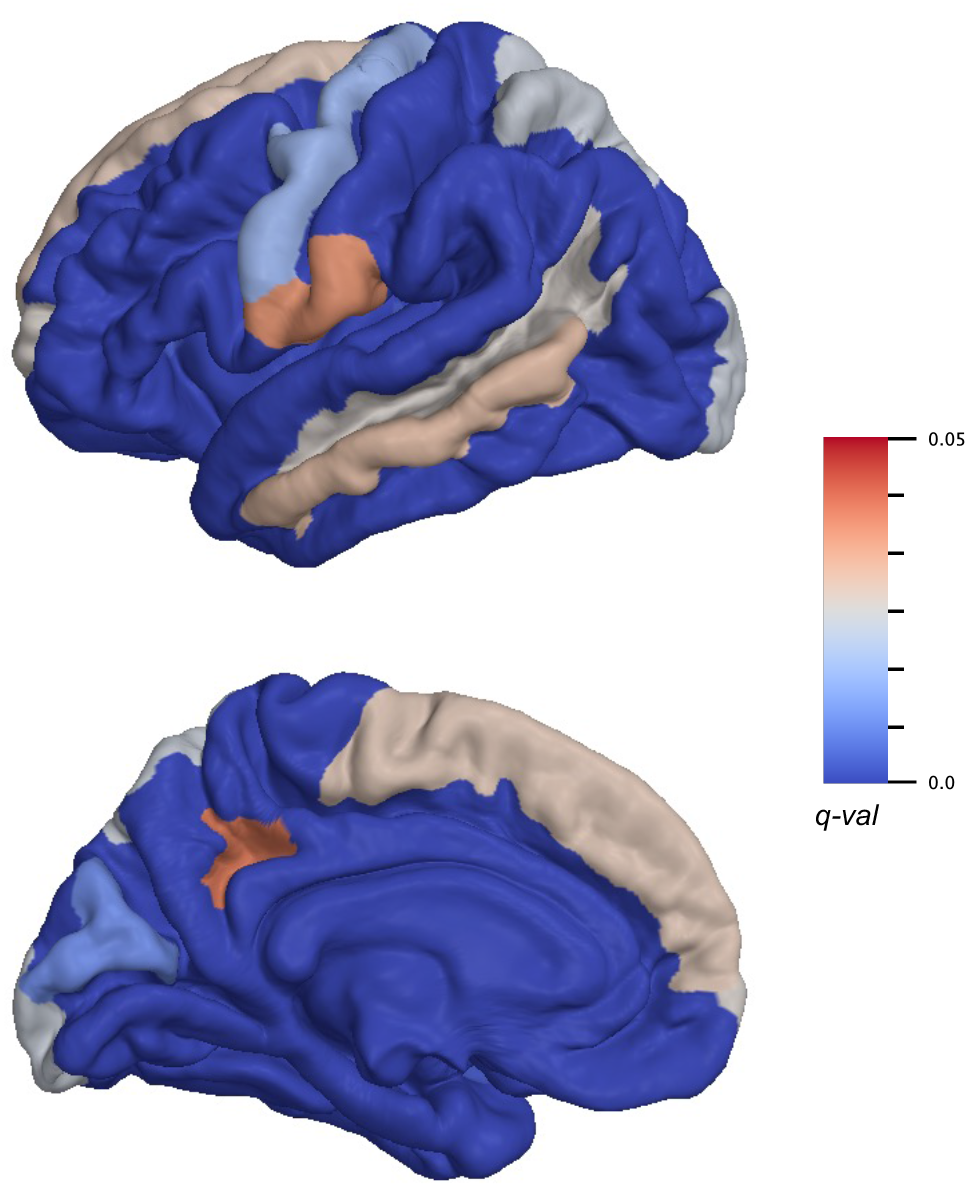
Brain regions in which cortical thickness is significantly associated to depression symptoms. Color bar indicates q-values. As results consider the two hemispheres together, only the left hemisphere is shown for visualization purposes.

#### 3.1.3. Somatization

Putamen (*q*=0.02), amygdala, hippocampus and thalamus volumes were associated with the somatization trait in patients with mTBI (all *q*<0.005). As for the WM volume, significant associations were seen in the fusiform, enthorinal, inferiortemporal, parahippocampal, precentral, paracentral, postcentral, posterior cingulate, supramarginal and superior-frontal areas (*q*<0.05). A significant association was found with the perivascular component in the fusiform, post-central and posterior cingulate areas, and overall WM volume (Figure 4). As for the tract morphological properties, the volume and length of the corticospinal tract and the anterior thalamic radiation were predictors of somatization (*q*<0.05) (Figure 1 & 5). A significant negative relationship was found with the MD of the uncinate fasciculus, inferior-fronto-orbital fasciculus, corticospinal tract, posterior arcuate, superior cerebellar tract, second branch of superior longitudinal fasciculus, fronto-anterior thalamic tract, superior thalamic radiation, middle longitudinal fasciculus, arcuate fasciculus, and anterior corona. For the last three tracts, a negative association was also seen with RD (Figure 5-6). Scatterplots of all statistically significant associations are shown in the Supplementary Materials.

**Figure 4:**
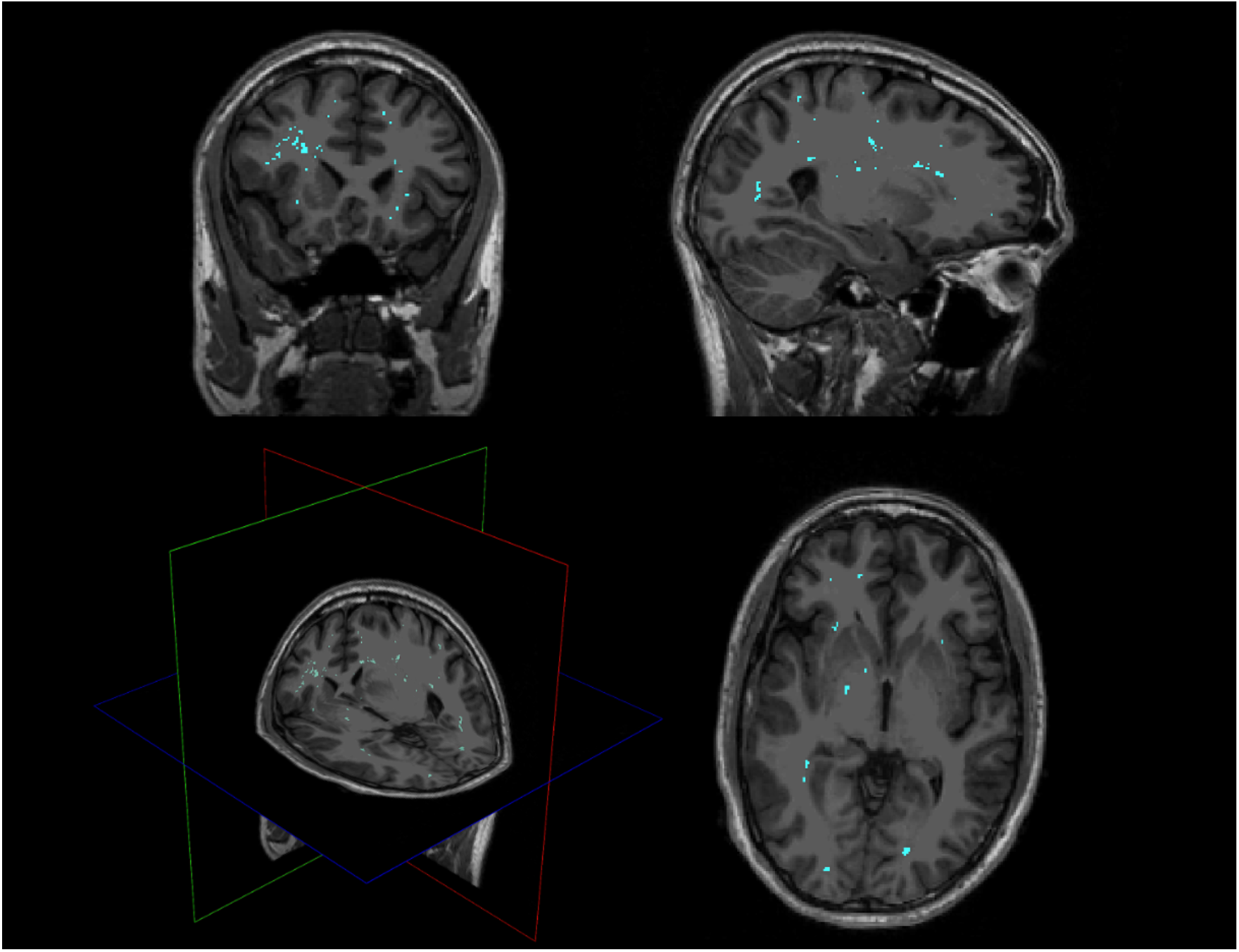
Perivascular space map on a single subject, shown on 3D plane, axial, sagittal and coronal views. Somatization symptoms were found to be significantly associated with global PVS volume in white matter, as well as locally in posterior cingulate, fusiform area and postcentral areas.

**Figure 5:**
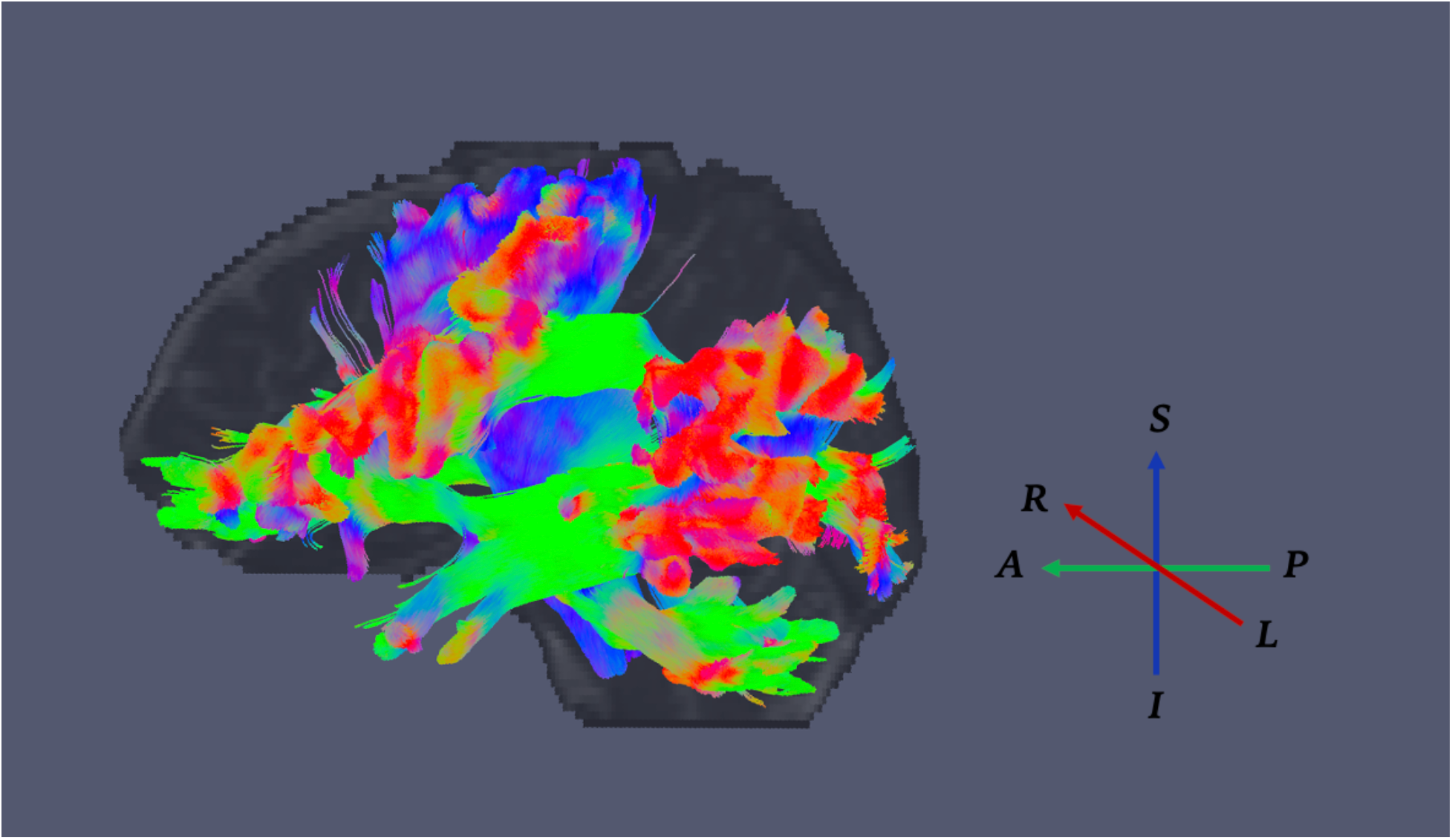
Visual representation of white matter tracts that were significantly different in diffusivity metrics (mean and radial diffusivity, MD and RD) when considering somatization symptoms. Arrows indicate the RGB color system of diffusion directions (A= Anterior, P=Posterior, S=Superior, I=Inferior, L=Left, R=Right).

**Figure 6:**
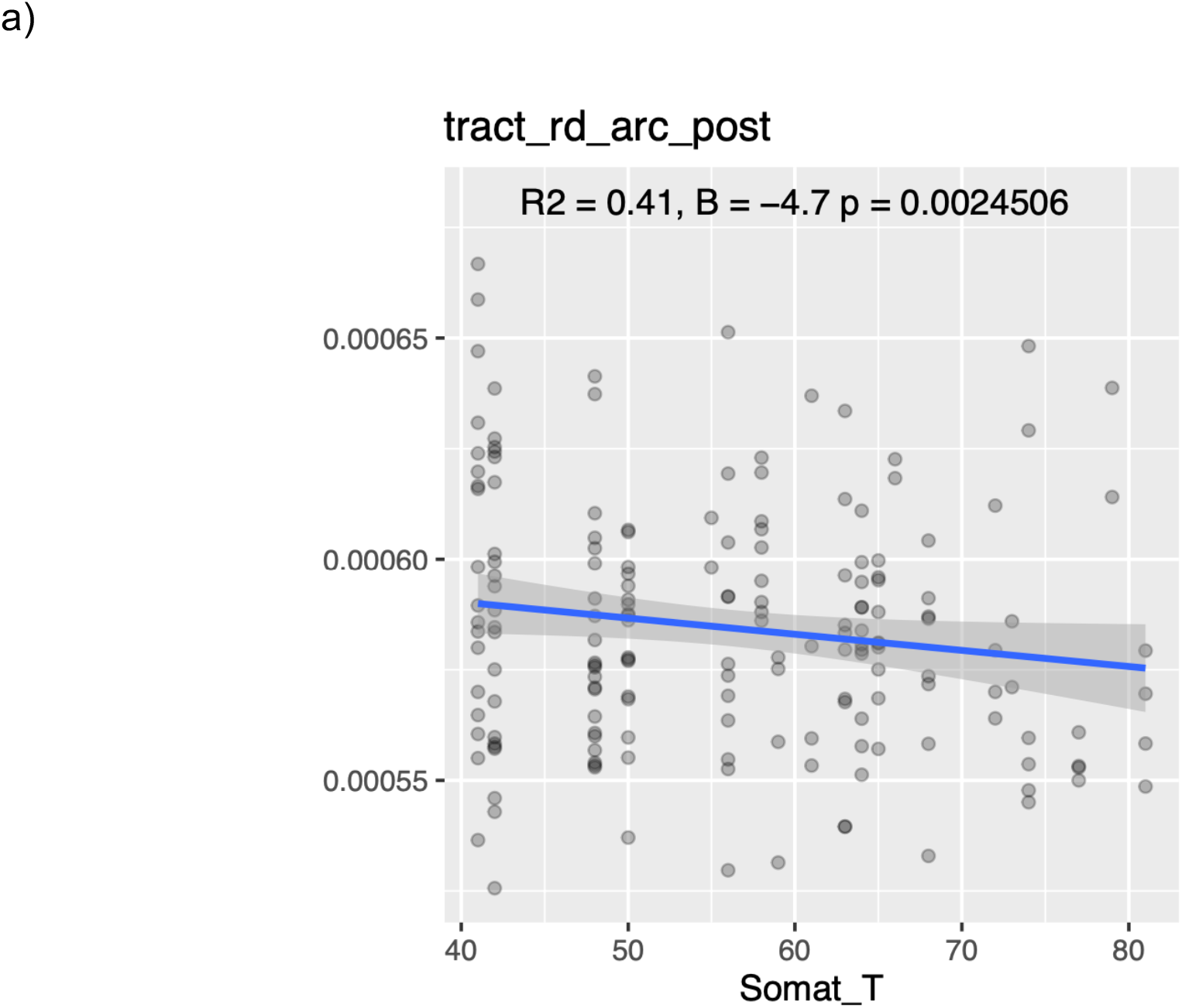
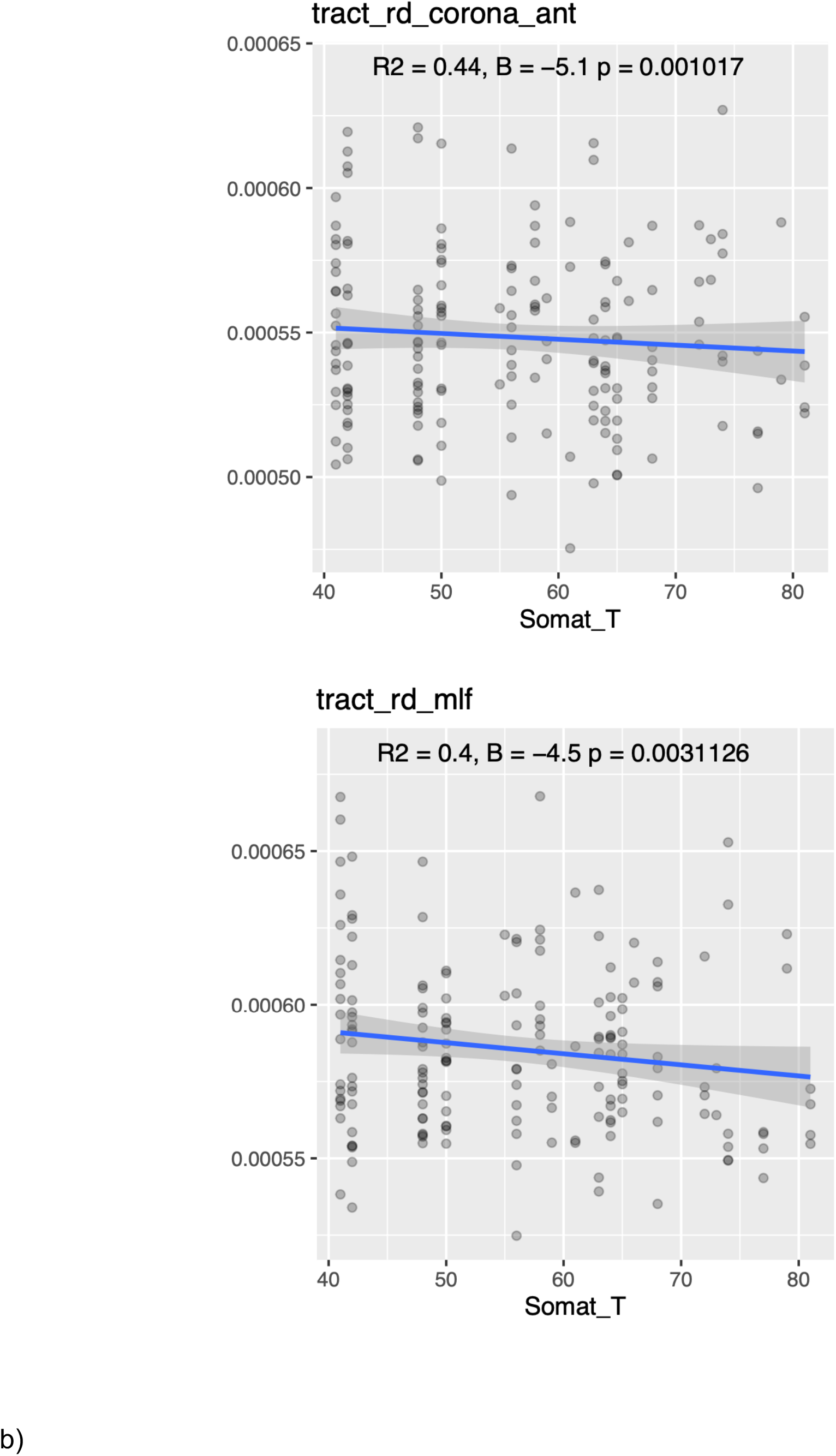
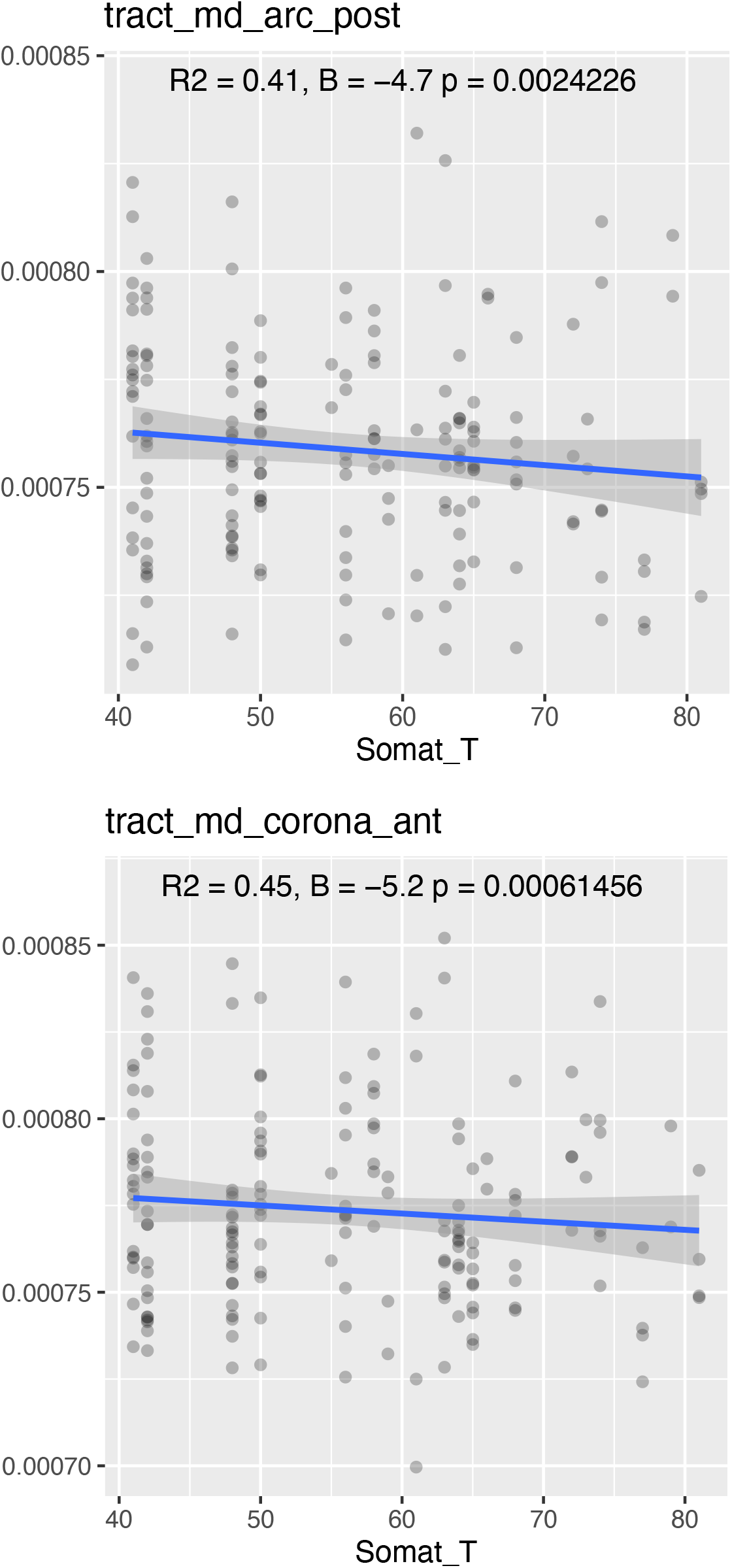
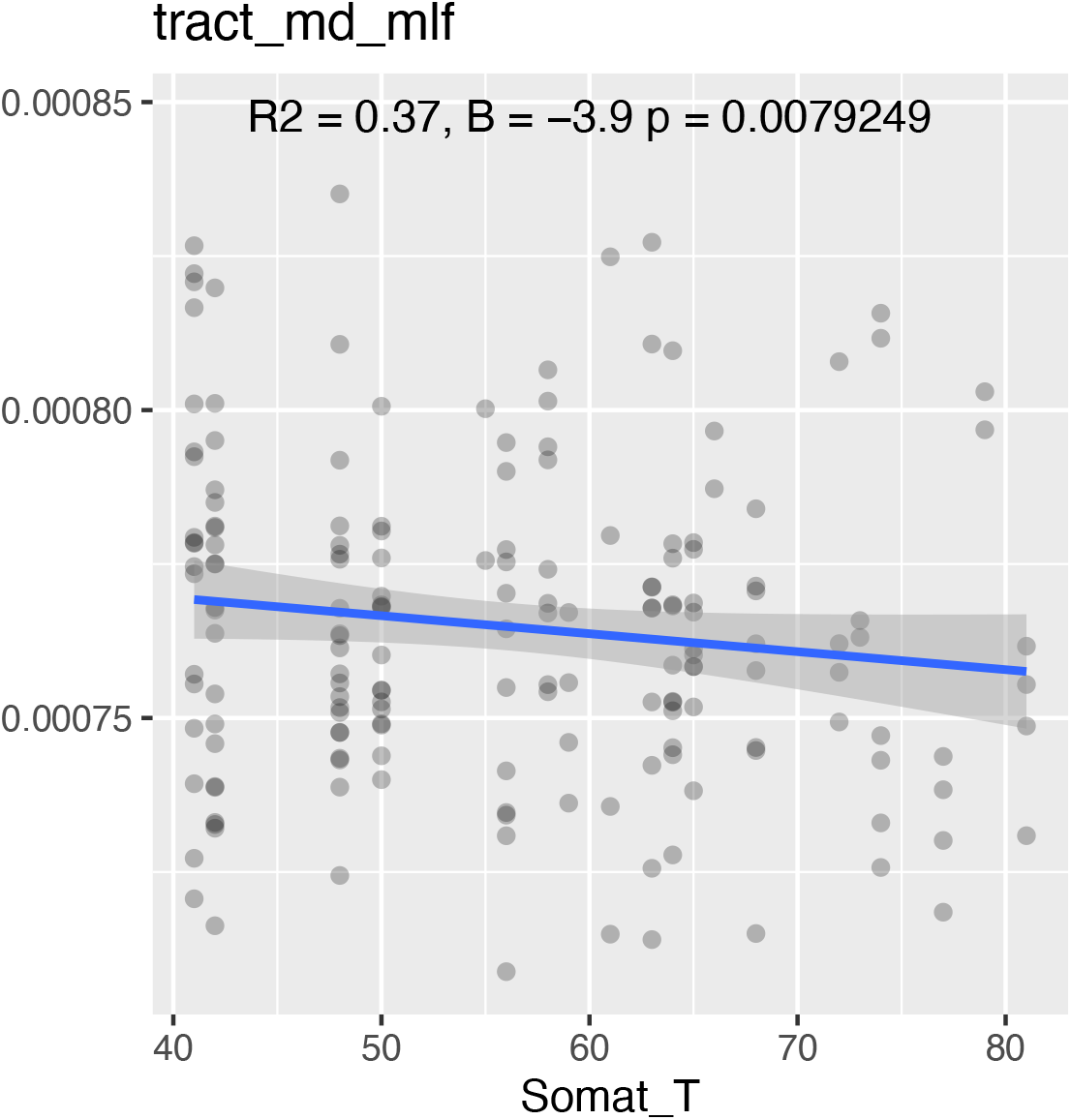
Scatterplots indicating a significant negative relationship of radial diffusivity (RD, a) and mean diffusivity (MD, b) with somatization symptoms in the posterior arcuate, anterior corona and middle longitudinal fasciculus. R-squared, beta and p-values are also indicated.

### 3.2. Life Satisfaction

Uncorrected *p*-values showed significant relationships with diffusivity measures and tract geometrical features of the cingulum, SLF-I, fornix, uncinate and arcuate fasciculus. Associations were found with insular perivascular spaces and cortical thickness, but they didn’t survive FDR correction. Uncorrected *p*-values, R^2^ and *β* values of all the brain regions predicting life satisfactory scores are reported in the Supplementary Material (Table S4 shows the two hemispheres together, in Table S5 results are reported for the two hemispheres separately).

## 4. Discussion

This study analyzed the relationship between structural imaging biomarkers and the development of depression, anxiety and somatization symptoms six months after mild TBI events. Findings showed the cortical thickness, diffusivity properties and perivascular components of brain regions predicting psychological distress are mostly involved in negative affective bias, emotional recall of memories and sensory-motor functions.

### 4.1. Somatization

A relationship between PVSs and development of somatization was found in the fusiform, post-central and posterior cingulate areas. This is the first time that PVS abnormalities have been reported to be predictor of somatization symptoms in mTBI patients. At a physiological level, the cognitive problems seen in mTBI patients, especially mnemonic and emotional domain, are to be associated to alteration of the glymphatic system functionality [49]; we would expect accompanying changes in the perivascular spaces, as they play a crucial role in the brain clearance activity. PVS enlargements result in a decreased flow of soluble waste, promoting the accumulation of neurotoxic molecules, such as beta-amyloid. Somatization is defined as the expression of mental distress as physical symptoms [50]. Lifestyle choices and physiological factors were previously found to trigger PVS structural alteration: for instance, higher body mass index potentially causes an increase of somatization symptoms severity [51] and PVS enlargement in white matter [52]. Likewise, poor quality of sleep and insomnia are important factors mediating such relationship, as just one night of deprived sleep has been shown to affect significantly perivascular space structure and function [53] [54]. Future studies will help understand the deeper relationship between the alteration of PVS anatomy and the onset of psychological distress symptoms.

The gray and white matter regions found to be significant predictors were previously shown to be structurally altered in first episode, drug-naïve patients with somatization disorder (SD) [55], reflecting changes in resting-state connectivity in the cortico-limbic- cerebellar circuit in patients with SD. The fusiform gyrus is part of the fusiform-amygdala circuit, and it is involved in social anxiety, avoidance, and sensitivity to punishment [56]. The posterior cingulate and post-central areas are key regions involved in basic emotion regulation processing, being part of the fronto-limbic and social networks [57]. Several studies have reported structural volume decreases in these areas in patients with somatization symptoms, related to higher scores of somatic symptoms [58] [59].

Results showed an association between increased amygdala thickness and psychological distress symptoms post TBI. This confirms previous findings that link amygdala increased connectivity and activation to impairment in memory, attention and learning on a cognitive level, as well as to depression, anxiety and fear-related behaviors on the emotional domain [60].

### 4.2. Depression

Depressive symptoms are among the most frequent psychiatric conditions post-mTBI [61], with a range between 10% to 77% among patients. Depression after mild TBI contributes to the exacerbation of post-concussive symptoms, such as dizziness, headache, sleep disorders, and increases in aggressive behavior and suicidal thoughts [61]. Neurochemical disruptions underlie post-TBI depressive symptoms, with chronic cholinergic deficits and dysfunction in dopaminergic, noradrenergic, and serotonergic systems all contributing [62]. Neurophysiological dysfunction and the severity of symptoms varies among patients and is influenced by the patient’s medical history (such as mood disorders, alcohol, or drug abuse), as well as by post-injury factors (for instance, lack of social supports or frustration in dealing with physical complications).

The internal capsule was found to be a significant predictor of both anxiety and depression symptoms in people with mTBI. The first study about diffuse axonal injury (DAI) on patients with mTBI [63] showed FA changes in the internal capsule, external capsule, and corpus callosum immediately after the traumatic event; our results suggest such changes can persist overtime. The diffusivity metric alterations underlie DAI, a form of TBI characterized by axon shearing at the junction of gray and white matter, leading to microstructural damage of fiber tract [64] and changes in the water molecule diffusion along the axons. In a group of mTBI patients following motor vehicle accident, diffusivity alterations were associated with worse processing speed and working memory indices [65], giving more insight on the neurobiological disruptions that lead to cognitive impairment caused by brain injuries. The internal capsule is one of the more commonly reported predictors of post-injury depression, together with the corpus callosum, anterior and posterior corona radiata, anterior and posterior thalamic radiations and corticospinal tract [66]. In our study, FA decreases and MD increases of these WM tracts were associated with anxiety, depression, and somatization symptoms altogether. Such regions are part of circuits involved in sleep-wake regulation, information processing, attention, executive function, and emotion regulation, all of which are shown to be impaired after mTBI.

### 4.3. Anxiety

Post-TBI anxiety disorder has a prevalence rate of up to 70%, [67], and can manifest in different forms across patients (i.e. anxiety symptoms, post-traumatic stress disorder, obsessive-compulsive disorder, panic disorder, and social anxiety disorder). Previous studies have found associations between anxiety and brain areas belonging to the fronto-limbic and corticostriatal pathways, such as the hippocampus, dorsolateral prefrontal and orbital frontal cortices, amygdala, and basal ganglia [68] [69] [70].

### 4.4. Post-injury recovery

In this study, we considered the relationship between MRI-based biomarkers and the onset of psychological distress after 6 months from injury. In this timeframe, the brain is subject to high plasticity levels that influence recovery stages. Depending on the injury severity and post-accident therapy, the brain can recover its functionality, thereby avoiding permanent impairment of cognitive abilities. A few studies showed a cognitive recovery within the first 6 months [5] [71] [72], whereas another study suggested that full or partial recovery will not always occur within such a period of time [73]. Recovery time varies highly among people, influenced by demographic and lifestyle factors such as age, educational level, and previous medical issues [73] [74] [75]. This highlights how structural alterations at 6 months following TBI need to be considered as a candidate biomarker and target for therapeutical interventions.

The findings of this study provide useful information regarding the anatomical areas targeted in mTBI. To localize the effects on psychological symptoms, we considered bilateral measures (obtained by combining the two hemispheres together) as well as hemispherical lateralization. The lateralized brain measures that were significantly associated with psychological distress symptoms confirm findings of a previous study looking at changes of brain connectivity in TBI patients [76]. They showed alterations in structural connectivity in the left thalamus, where we found significant relationships with somatization, and in the right postcentral gyrus, significantly associated with somatization symptoms when considering PVS fraction. This suggests that changes in the different structural compartments and network reorganization following TBI may exacerbate somatization symptoms, as these brain areas are involved in sensation and limbic functions.

To our knowledge, no prior study has yet investigated the association of perivascular space and psychological distress in mTBI patients. An alteration in the PVS structure can lead to disruption in the blood brain barrier (BBB) and alteration in the cerebrovascular physiological flow, promoting deposits of neurotoxic molecules.

Anatomically, PVS enlargements can lead to failure in the BBB and the vascular component supplying such region, namely the anterior and middle cerebral arteries [77]. A recent paper quantified BBB permeability increase in animal models with repeated mild head injuries by using dynamic contrast enhanced (DCE)-MRI. They found that, at first head impact, the BBB permeability increased particularly in the somatosensory and frontal cortices [78]. The alteration in BBB integrity caused by brain injury can lead to long-term inflammatory states and culminate in permanent changes in brain homeostasis [79].

### 4.5. Limitations

In the present study we investigated the relationship of psychological distress symptoms and structural imaging biomarkers cross-sectionally. We were able to collect BSI information at 6 months only and not further timepoints. A longitudinal component beyond 6 months could be helpful in monitoring the symptom severity after the acute and semi-acute post-injury stages and infer on the recovery speed. The cross-sectional approach to the analysis also explains the relatively small sample size, represented by participants who had completed the demographic information and self-reported psychological measures in the TRACK-TBI cohort, and considering only MRI and DWI with the best quality data. Another potential limitation is considering only structural MRI and not other neuroimaging techniques. Even if we present an exhaustive analysis on the diffusion, volumetric, and perivascular components that predict psychological consequences in TBI patients, a multi-modal approach can help in further understanding the variability in the severity of psychological and cognitive outcome among TBI patients. It is worth pointing out that the diffusion measures and the tract geometrical features computed for the analysis have limited reproducibility across fiber bundle segmentation approaches from different research groups, as shown by a recent study [80]. The high variability in WM tract reconstruction influences metrics quantification and, therefore, is an important factor to consider with the interpretation of microstructural changes in clinical populations in conditions when data are pooled across studies.

### 4.6. Conclusions

Our findings not only confirm previous results on the volumetric and diffusion-based associations to depressive and anxiety symptoms but reveal a further significant relationship between the PVS anatomy and somatization disorder. This possibly reflects abnormalities in the brain glymphatic system and BBB integrity, that play a crucial role in brain waste clearance. Alterations of these physiological processes can trigger a cascade of events responsible for neurological disorders development and cognitive decline [81]. Results confirm that the brain is affected by mTBI episodes on different structural compartments, leading to accelerated neurodegenerative and neuroinflammatory processes, if not monitored over time. The role of efficient therapeutical interventions is therefore crucial to prevent the exacerbation of the psychological symptoms and cognitive decline seen already at mild stages of brain injuries, and the effects reported here may provide a possible avenue for treatment evaluation in future studies.

*TRACK-TBI Investigators: Shelly R. Cooper, BA (Department of Neurological Surgery, University of California, San Francisco, San Francisco, CA); (Kristen Dams-O’Connor, PhD (Department of Rehabilitation Medicine, Icahn School of Medicine at Mount Sinai, New York, NY); Wayne A. Gordon, PhD (Department of Rehabilitation Medicine, Icahn School of Medicine at Mount Sinai, New York, NY); Andrew I.R. Maas, MD, PhD (Department of Neurological Surgery, University Hospital Antwerp, Antwerp, Belgium); David K. Menon, MD, PhD (Departments of Anaesthesia and Neurocritical Care, University of Cambridge, Cambridge, United Kingdom); Pratik Mukherjee, MD, PhD (Department of Radiology, University of California, San Francisco, San Francisco, CA); Ava M. Puccio, RN, PhD (Department of Neurological Surgery, University of Pittsburgh Medical Center, Pittsburgh, PA); Mary J. Vassar, RN, MS (Department of Neurological Surgery, University of California, San Francisco, San Francisco, CA); John K. Yue, MD (Department of Neurological Surgery, University of California, San Francisco, San Francisco, CA); and Esther L. Yuh, MD, PhD (Department of Radiology, University of California, San Francisco, San Francisco, CA).

## Data Availability

All data produced in the present study are available upon reasonable request to the authors

https://fitbir.nih.gov

## Acknowledgments

This work is supported directly by the National Institutes of Health (NIH) grant number 5R01NS100973-05. The image computing resources provided by the Laboratory of Neuro Imaging Resource (LONIR) at USC are supported in part by NIH grant number P41EB015922. Author RPC is supported in part by grant number 2020-225670 from the Chan Zuckerberg Initiative DAF, an advised fund of Silicon Valley Community Foundation.

Data and research tools used in the preparation of this article reside in and were analyzed using the Department of Defense (DOD) and National Institutes of Health (NIH)-supported Federal Interagency Traumatic Brain Injury Research Informatics System (FITBIR). FITBIR is a collaborative biomedical informatics system created by DOD and NIH to provide a national resource to support and accelerate research in traumatic brain injury. Dataset identifier: [doi:10.23718/FITBIR/1419836]. This manuscript reflects the views of the authors and does not reflect the opinions or views of the DOD or NIH.

## References

1. Khellaf, A., Khan, D. Z., & Helmy, A. (2019). Recent advances in traumatic brain injury. Journal of Neurology, 266(11), 2878–2889. doi:10.1007/s00415-019-09541-4 [doi]

2. Najem, D., Rennie, K., Ribecco-Lutkiewicz, M., Ly, D., Haukenfrers, J., Liu, Q., . . . Bani-Yaghoub, M. (2018). Traumatic brain injury: Classification, models, and markers. Biochemistry and Cell Biology = Biochimie Et Biologie Cellulaire, 96(4), 391–406. doi:10.1139/bcb-2016-0160 [doi]

3. Iverson, G. L. (2005). Outcome from mild traumatic brain injury. Current Opinion in Psychiatry, 18(3).

4. Vos, P. E., Alekseenko, Y., Battistin, L., Ehler, E., Gerstenbrand, F., Muresanu, D. F., . . . von Wild, K. (2012). Mild traumatic brain injury. European Journal of Neurology, 19(2), 191–198. doi:https://doi.org/10.1111/j.1468-1331.2011.03581.x

5. Stulemeijer, M., van der Werf, S., Borm, G. F., & Vos, P. E. (2008). Early prediction of favourable recovery 6 months after mild traumatic brain injury. J Neurol Neurosurg Psychiatry, 79(8), 936. doi:10.1136/jnnp.2007.131250

6. Quatman-Yates, C., Cupp, A., Gunsch, C., Haley, T., Vaculik, S., & Kujawa, D. (2016). Physical rehabilitation interventions for post-mTBI symptoms lasting greater than 2 weeks: Systematic review. Physical Therapy, 96(11), 1753–1763. doi:10.2522/ptj.20150557

7. Ridner, S. H. (2004). Psychological distress: Concept analysis. Journal of Advanced Nursing, 45(5), 536–545. doi:https://doi.org/10.1046/j.1365-2648.2003.02938.x

8. McCrea, M., Guskiewicz, K. M., Marshall, S. W., Barr, W., Randolph, C., Cantu, R. C., . . . Kelly, J. P. (2003). Acute effects and recovery time following concussion in collegiate football players: The NCAA concussion study. Jama, 290(19), 2556–2563. doi:290/19/2556 [pii]

9. Jorge, R. E., Robinson, R. G., Moser, D., Tateno, A., Crespo-Facorro, B., & Arndt, S. (2004). Major depression following traumatic brain injury. Archives of General Psychiatry, 61(1), 42–50. doi:10.1001/archpsyc.61.1.42

10. Leong Bin Abdullah, Mohammad, Farris Iman, Ng, Y. P., & Sidi, H. B. (2018). Depression and anxiety among traumatic brain injury patients in malaysia. Asian Journal of Psychiatry, 37, 67–70. doi:https://doi.org/10.1016/j.ajp.2018.08.017

11. Hellewell, S. C., Beaton, C. S., Welton, T., & Grieve, S. M. (2020). Characterizing the risk of depression following mild traumatic brain injury: A meta-analysis of the literature comparing chronic mTBI to non-mTBI populations. Frontiers in Neurology, 11, 350. doi:10.3389/fneur.2020.00350

12. van der Naalt, J., Timmerman, M. E., de Koning, M.,E., van der Horn, H., J., Scheenen, M. E., Jacobs, B., . . . Spikman, J. M. (2017). Early predictors of outcome after mild traumatic brain injury (UPFRONT): An observational cohort study. The Lancet Neurology, 16(7), 532–540. doi:https://doi.org/10.1016/S1474-4422(17)30117-5

13. Bigler, E. D., Abildskov, T. J., Goodrich-Hunsaker, N., Black, G., Christensen, Z. P., Huff, T., . . . Max, J. E. (2016). Structural neuroimaging findings in mild traumatic brain injury. Sports Medicine and Arthroscopy Review, 24(3), e42–e52. doi:10.1097/JSA.0000000000000119

14. Marum, G., Clench-Aas, J., Nes, R. B., & Raanaas, R. K. (2014). The relationship between negative life events, psychological distress and life satisfaction: A population-based study. Quality of Life Research, 23(2), 601–611. doi:10.1007/s11136-013-0512-8

15. Juengst, S. B., Adams, L. M., Bogner, J. A., Arenth, P. M., O’Neil-Pirozzi, T. M., Dreer, L. E., . . . Wagner, A. K. (2015). Trajectories of life satisfaction after traumatic brain injury: Influence of life roles, age, cognitive disability, and depressive symptoms. Rehabilitation Psychology, 60(4), 353–364. doi:10.1037/rep0000056 [doi]

16. Reith, F. C. M., Van den Brande, R., Synnot, A., Gruen, R., & Maas, A. I. R. (2016). The reliability of the glasgow coma scale: A systematic review. Intensive Care Medicine, 42(1), 3–15. doi:10.1007/s00134-015-4124-3

17. Franke, G. H., Jaeger, S., Glaesmer, H., Barkmann, C., Petrowski, K., & Braehler, E. (2017). Psychometric analysis of the brief symptom inventory 18 (BSI-18) in a representative german sample. BMC Medical Research Methodology, 17(1), 14. doi:10.1186/s12874-016-0283-3

18. Derogatis, L. R. (1993). BSI Brief Symptom Inventory. Administration, Scoring, and Procedures Manual (4th Ed.). Minneapolis, MN: National Computer Systems.

19. Meijer, R. R., de Vries, R. M., & van Bruggen, V. (2011). An evaluation of the brief symptom inventory-18 using item response theory: Which items are most strongly related to psychological distress? Psychological Assessment, 23(1), 193–202. doi:10.1037/a0021292 [doi]

20. Recklitis, C. J., Blackmon, J. E., & Chang, G. (2017). Validity of the brief symptom inventory-18 (BSI-18) for identifying depression and anxiety in young adult cancer survivors: Comparison with a structured clinical diagnostic interview. Psychological Assessment, 29(10), 1189–1200. doi:10.1037/pas0000427

21. Shin, D. C., & Johnson, D. M. (1978). Avowed happiness as an overall assessment of the quality of life. Social Indicators Research, 5(1), 475–492. doi:10.1007/BF00352944

22. Diener, E., Emmons, R. A., Larsen, R. J., & Griffin, S. (1985). The Satisfaction With Life Scale. Journal of Personality Assessment, 49(1), 71–75. https://doi.org/10.1207/s15327752jpa4901_13

23. Pavot, W., & Diener, E. (1993). Review of the Satisfaction With Life Scale. Psychological Assessment, 5(2), 164–172

24. Yuh, E. L., Jain, S., Sun, X., Pisică, D., Harris, M. H., Taylor, S. R., . . . TRACK-TBI Investigators for the CENTER-TBI Investigators. (2021). Pathological computed tomography features associated with adverse outcomes after mild traumatic brain injury: A TRACK-TBI study with external validation in CENTER-TBI. JAMA Neurology, doi:10.1001/jamaneurol.2021.2120

25. Jenkinson, M., Beckmann, C. F., Behrens, T. E., Woolrich, M. W., & Smith, S. M. (2012). Fsl. Neuroimage, 62(2), 782–790.

26. Avants, B. B., Tustison, N., & Song, G. (2009). Advanced normalization tools (ANTS). Insight j, 2(365), 1–35.

27. Cabeen, R. P., Laidlaw, D. H., & Toga, A. W. (2018). Quantitative imaging toolkit: Software for interactive 3D visualization, data exploration, and computational analysis of neuroimaging datasets. ISMRM-ESMRMB Abstracts, , 12-14.

28. Manjón, J. V., Coupé, P., Martí-Bonmatí, L., Collins, D. L., & Robles, M. (2010). Adaptive non-local means denoising of MR images with spatially varying noise levels. Journal of Magnetic Resonance Imaging, 31(1), 192–203.

29. Andersson, J. L. R., & Sotiropoulos, S. N. (2016). An integrated approach to correction for off-resonance effects and subject movement in diffusion MR imaging. NeuroImage, 125, 1063–1078. doi:S1053-8119(15)00920-9 [pii]

30. Bastiani, M., Cottaar, M., Fitzgibbon, S. P., Suri, S., Alfaro-Almagro, F., Sotiropoulos, S. N., . . . Andersson, J. L. R. (2019). Automated quality control for within and between studies diffusion MRI data using a non-parametric framework for movement and distortion correction. NeuroImage, 184, 801–812. doi:S1053- 8119(18)31945-1 [pii]

31. Smith, S. M. (2002). Fast robust automated brain extraction. Human brain mapping, 17(3), 143–155.

32. Veraart, J., Sijbers, J., Sunaert, S., Leemans, A., & Jeurissen, B. (2013). Weighted linear least squares estimation of diffusion MRI parameters: strengths, limitations, and pitfalls. Neuroimage, 81, 335–346.

33. Alexander, A. L., Lee, J. E., Lazar, M., & Field, A. S. (2007). Diffusion tensor imaging of the brain. Neurotherapeutics, 4(3), 316–329.

34. Behrens, T. E., Berg, H. J., Jbabdi, S., Rushworth, M. F., & Woolrich, M. W. (2007). Probabilistic diffusion tractography with multiple fibre orientations: What can we gain?. Neuroimage, 34(1), 144–155.

35. Lynch, K. M., Cabeen, R. P., Toga, A. W., & Clark, K. A. (2020). Magnitude and timing of major white matter tract maturation from infancy through adolescence with NODDI. NeuroImage, 212, 116672.

36. Cabeen, R. P., Bastin, M. E., & Laidlaw, D. H. (2016). Kernel regression estimation of fiber orientation mixtures in diffusion MRI. Neuroimage, 127, 158–172.

37. Zhang, S., Peng, H., Dawe, R. J., & Arfanakis, K. (2011). Enhanced ICBM diffusion tensor template of the human brain. Neuroimage, 54(2), 974–984.

38. Catani, M., Dell’Acqua, F., Vergani, F., Malik, F., Hodge, H., Roy, P., et al. (2012). Short frontal lobe connections of the human brain. Cortex 48, 273–291. doi: 10.1016/j.cortex.2011.12.001

39. Mori, S., Oishi, K., Jiang, H., Jiang, L., Li, X., Akhter, K., … & Mazziotta, J. (2008). Stereotaxic white matter atlas based on diffusion tensor imaging in an ICBM template. Neuroimage, 40(2), 570–582.

40. Avants, B. B., Epstein, C. L., Grossman, M., & Gee, J. C. (2008). Symmetric diffeomorphic image registration with cross-correlation: evaluating automated labeling of elderly and neurodegenerative brain. Medical image analysis, 12(1), 26–41.

41. Cabeen, R. P., & Toga, A. W. (2020). Reinforcement Tractography: A Hybrid Approach for Robust Segmentation of Complex Fiber Bundles. In 2020 IEEE 17th International Symposium on Biomedical Imaging (ISBI) (pp. 999-1003). IEEE.

42. Fischl, B. (2012). FreeSurfer. Neuroimage, 62(2), 774–781.

43. Desikan, R. S., Ségonne, F., Fischl, B., Quinn, B. T., Dickerson, B. C., Blacker, D., … & Killiany, R. J. (2006). An automated labeling system for subdividing the human cerebral cortex on MRI scans into gyral based regions of interest. Neuroimage, 31(3), 968–980.

44. Sepehrband, F., Barisano, G., Sheikh-Bahaei, N., Cabeen, R. P., Choupan, J., Law, M., & Toga, A. W. (2019). Image processing approaches to enhance perivascular space visibility and quantification using MRI. Scientific Reports, 9(1), 12351. doi:10.1038/s41598-019-48910-x

45. Mori et al., MRI Atlas of Human White Matter. Elsevier, Amsterdam, The Netherlands (2005)

46. Wakana et al., Reproducibility of quantitative tractography methods applied to cerebral white matter. NeuroImage 36:630–644 (2007)

47. Hua et al., Tract probability maps in stereotaxic spaces: analysis of white matter anatomy and tract-specific quantification. NeuroImage, 39(1):336–347 (2008)

48. Benjamini, Y., Drai, D., Elmer, G., Kafkafi, N., & Golani, I. (2001). Controlling the false discovery rate in behavior genetics research. Behavioural brain research, 125(1-2), 279–284.

49. Scott, G., Ramlackhansingh, A. F., Edison, P., Hellyer, P., Cole, J., Veronese, M., . . . Sharp, D. J. (2016). Amyloid pathology and axonal injury after brain trauma. Neurology, 86(9), 821–828. doi:10.1212/WNL.0000000000002413 [doi]

50. Fu, X., Zhang, F., Liu, F., Yan, C., & Guo, W. (2019). Editorial: Brain and somatization symptoms in psychiatric disorders. Frontiers in Psychiatry, 10, 146. doi:10.3389/fpsyt.2019.00146

51. Kim, H. J., Kim, H. R., Jin, J., Han, D. H., & Kim, S. M. (2021). Body mass index and somatic symptom severity in patients with somatic symptom disorder: The mediating role of working memory. Clinical Psychopharmacology and Neuroscience : The Official Scientific Journal of the Korean College of Neuropsychopharmacology, 19(2), 361–366. doi:10.9758/cpn.2021.19.2.361

52. Barisano, G., Sheikh-Bahaei, N., Law, M., Toga, A. W., & Sepehrband, F. (2020). Body mass index, time of day, and genetics affect perivascular spaces in the white matter. J Cereb Blood Flow Metab, , 0271678X20972856. doi:10.1177/0271678X20972856

53. Zhang, J., Lam, S. P., Li, S. X., Tang, N. L., Yu, M. W. M., Li, A. M., & Wing, Y. K. (2012). Insomnia, sleep quality, pain, and somatic symptoms: Sex differences and shared genetic components. Pain, 153(3), 666–673. doi:10.1016/j.pain.2011.12.003 [doi]

54. Shokri-Kojori, E., Wang, G. J., Wiers, C. E., Demiral, S. B., Guo, M., Kim, S. W., . . . Volkow, N. D. (2018). Β-amyloid accumulation in the human brain after one night of sleep deprivation. Proceedings of the National Academy of Sciences of the United States of America, 115(17), 4483–4488. doi:10.1073/pnas.1721694115 [doi]

55. Li, R., Liu, F., Su, Q., Zhang, Z., Zhao, J., Wang, Y., Wu, R., Zhao, J., Guo, W. (2018). Bidirectional causal connectivity in the cortico-limbic-cerebellar circuit related to structural alterations in first-episode, drug-naive somatization disorder. Frontiers in Psychiatry, 9, 162.

56. Pujol, J., Harrison, B. J., Ortiz, H., Deus, J., Soriano-Mas, C., López-Solà, M., . . . Cardoner, N. (2009). Influence of the fusiform gyrus on amygdala response to emotional faces in the non-clinical range of social anxiety. Psychological Medicine, 39(7), 1177–1187. doi:10.1017/S003329170800500X

57. Li, X., Zhang, M., Li, K., Zou, F., Wang, Y., Wu, X., & Zhang, H. (2019). The altered somatic brain network in state anxiety. Frontiers in Psychiatry, 10, 465. doi:10.3389/fpsyt.2019.00465

58. McLaren, M. E., Szymkowicz, S. M., O’Shea, A., Woods, A. J., Anton, S. D., & Dotson, V. M. (2016). Dimensions of depressive symptoms and cingulate volumes in older adults. Translational Psychiatry, 6(4), e788. doi:10.1038/tp.2016.49

59. Lemche, E., Giampietro, V. P., Brammer, M. J., Surguladze, S. A., Williams, S. C. R., & Phillips, M. L. (2013). Somatization severity associated with postero-medial complex structures. Scientific Reports, 3(1), 1032. doi:10.1038/srep01032

60. McCorkle, T. A., Barson, J. R., & Raghupathi, R. (2021). A role for the amygdala in impairments of affective behaviors following mild traumatic brain injury. Frontiers in Behavioral Neuroscience, 15, 601275. doi:10.3389/fnbeh.2021.601275

61. Levin, H. S., McCauley, S. R., Josic, C. P., Boake, C., Brown, S. A., Goodman, H. S., . . . Brundage, S. I. (2005). Predicting depression following mild traumatic brain injury. Archives of General Psychiatry, 62(5), 523–528. doi:10.1001/archpsyc.62.5.523

62. Silver, J. M., McAllister, T. W., & Arciniegas, D. B. (2009). Depression and cognitive complaints following mild traumatic brain injury. Ajp, 166(6), 653–661. doi:10.1176/appi.ajp.2009.08111676

63. Arfanakis, K., Haughton, V. M., Carew, J. D., Rogers, B. P., Dempsey, R. J., & Meyerand, M. E. (2002). Diffusion tensor MR imaging in diffuse axonal injury. AJNR.American Journal of Neuroradiology, 23(5), 794–802.

64. Jolly, A. E., Bălăeţ, M., Azor, A., Friedland, D., Sandrone, S., Graham, N. S. N., . . . Sharp, D. J. (2021). Detecting axonal injury in individual patients after traumatic brain injury. Brain, 144(1), 92–113. doi:10.1093/brain/awaa372

65. Xiong, K., Zhu, Y., Zhang, Y., Yin, Z., Zhang, J., Qiu, M., & Zhang, W. (2014). White matter integrity and cognition in mild traumatic brain injury following motor vehicle accident. Brain Research, 1591, 86–92. doi:https://doi.org/10.1016/j.brainres.2014.10.030

66. Raikes, A. C., Bajaj, S., Dailey, N. S., Smith, R. S., Alkozei, A., Satterfield, B. C., & Killgore, W. D. S. (2018). Diffusion tensor imaging (DTI) correlates of self-reported sleep quality and depression following mild traumatic brain injury. Frontiers in Neurology, 9, 468.

67. Mallya, S., Sutherland, J., Pongracic, S., Mainland, B., & Ornstein, T. J. (2015). The manifestation of anxiety disorders after traumatic brain injury: A review. Journal of Neurotrauma, 32(7), 411–421. doi:10.1089/neu.2014.3504

68. Tromp, D. P. M., Grupe, D. W., Oathes, D. J., McFarlin, D. R., Hernandez, P. J., Kral, T. R. A., . . . Nitschke, J. B. (2012). Reduced structural connectivity of a major frontolimbic pathway in generalized anxiety disorder. Archives of General Psychiatry, 69(9), 925–934. doi:10.1001/archgenpsychiatry.2011.2178

69. Macpherson, T., & Hikida, T. (2019). Role of basal ganglia neurocircuitry in the pathology of psychiatric disorders. Psychiatry and Clinical Neurosciences, 73(6), 289–301. doi:https://doi.org/10.1111/pcn.12830

70. Madonna, D., Delvecchio, G., Soares, J. C., & Brambilla, P. (2019). Structural and functional neuroimaging studies in generalized anxiety disorder: A systematic review. Revista Brasileira De Psiquiatria (Sao Paulo, Brazil : 1999), 41(4), 336–362. doi:10.1590/1516-4446-2018-0108

71. Novack, T. A., Alderson, A. L., Bush, B. A., Meythaler, J. M., & Canupp, K. (2000). Cognitive and functional recovery at 6 and 12 months post-TBI. Brain Injury, 14(11), 987–996. doi:10.1080/02699050050191922 [doi]

72. Bharath, R. D., Munivenkatappa, A., Gohel, S., Panda, R., Saini, J., Rajeswaran, J., . . . Biswal, B. B. (2015). Recovery of resting brain connectivity ensuing mild traumatic brain injury. Frontiers in Human Neuroscience, 9, 513. doi:10.3389/fnhum.2015.00513

73. Triebel, K. L., Martin, R. C., Novack, T. A., Dreer, L. E., Turner, C., Kennedy, R., & Marson, D. C. (2014). Recovery over 6 months of medical decision-making capacity after traumatic brain injury. Archives of Physical Medicine and Rehabilitation, 95(12), 2296–2303. doi:10.1016/j.apmr.2014.07.413

74. Mooney, G., Speed, J., & Sheppard, S. (2005). Factors related to recovery after mild traumatic brain injury. Null, 19(12), 975–987. doi:10.1080/02699050500110264

75. Yue, J. K., Levin, H. S., Suen, C. G., Morrissey, M. R., Runyon, S. J., Winkler, E. A., . . . TRACK-TBI Investigators. (2019). Age and sex-mediated differences in six-month outcomes after mild traumatic brain injury in young adults: A TRACK-TBI study. Neurological Research, 41(7), 609–623. doi:10.1080/01616412.2019.1602312

76. Caeyenberghs, K., Leemans, A., De Decker, C., Heitger, M., Drijkoningen, D., Linden, C. V., . . . Swinnen, S. P. (2012). Brain connectivity and postural control in young traumatic brain injury patients: A diffusion MRI based network analysis. NeuroImage.Clinical, 1(1), 106–115. doi:10.1016/j.nicl.2012.09.011 [doi]

77. Wardlaw JM. Blood-brain barrier and cerebral small vessel disease. J Neurol Sci. (2010) 299:66–71. doi: 10.1016/j.jns.2010.08.042

78. Leaston, J., Qiao, J., Harding, I. C., Kulkarni, P., Gharagouzloo, C., Ebong, E., & Ferris, C. F. (2021). Quantitative imaging of blood-brain barrier permeability following repetitive mild head impacts. Frontiers in Neurology, 12, 1654.

79. Li, W., Watts, L., Long, J., Zhou, W., Shen, Q., Jiang, Z., Li, Y. Duong, T. Q. (2016). Spatiotemporal changes in blood-brain barrier permeability, cerebral blood flow, T2 and diffusion following mild traumatic brain injury. Brain Research, 1646, 53–61.

80. Schilling, K. G., Rheault, F., Petit, L., Hansen, C. B., Nath, V., Yeh, F. C., … & Descoteaux, M. (2021). Tractography dissection variability: what happens when 42 groups dissect 14 white matter bundles on the same dataset?. NeuroImage, 243, 118502

81. Wu, Y., Wu, H., Guo, X., Pluimer, B., & Zhao, Z. (2020). Blood–Brain barrier dysfunction in mild traumatic brain injury: Evidence from preclinical murine models. Frontiers in Physiology, 11, 1030.

