## Supplementary Material for "Life after mild traumatic brain injury: Widespread structural brain changes associated with psychological distress revealed with multimodal magnetic resonance imaging"

Supplementary Methods

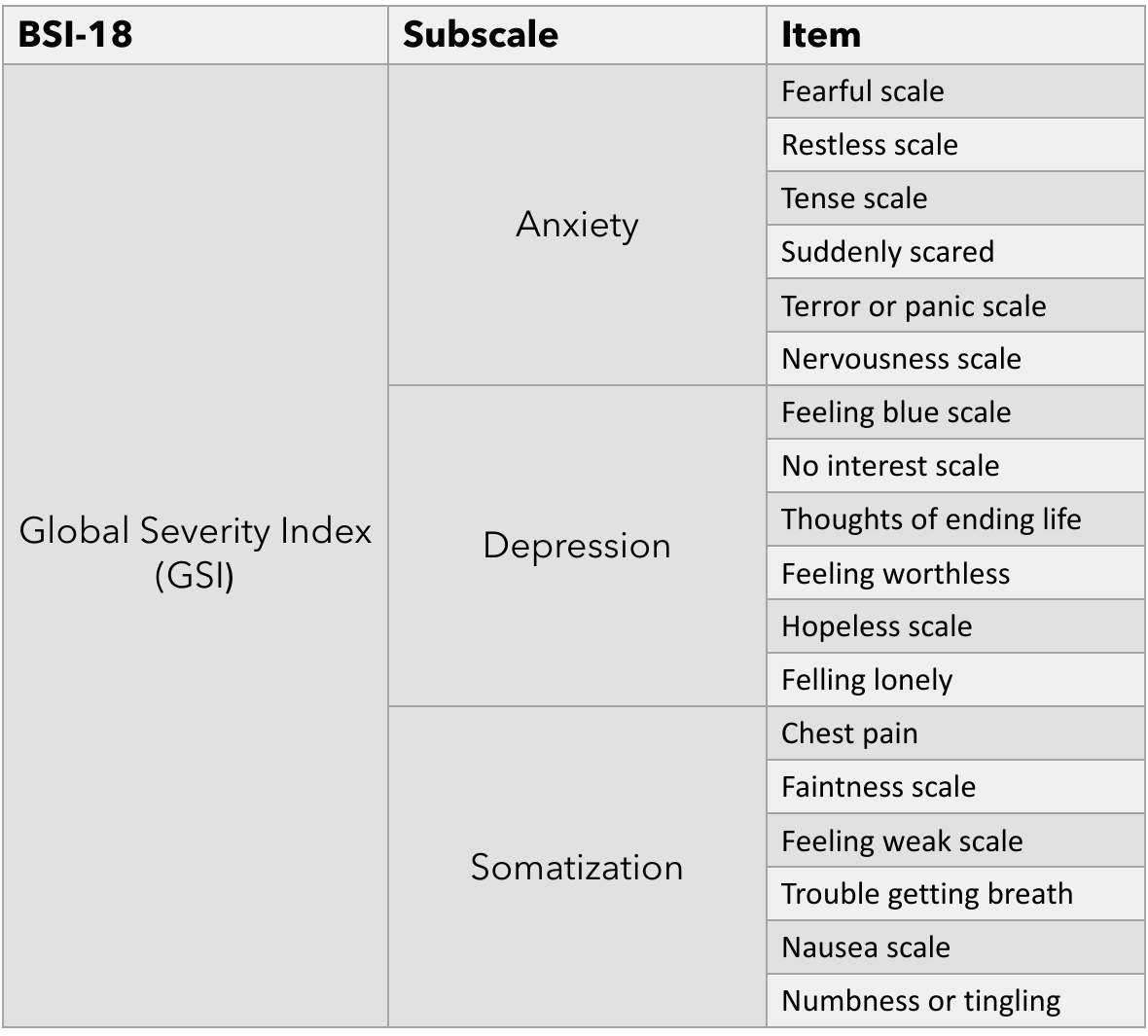

**Table S1:** List of the items composing the three subscales of BSI-18 used for the analysis.

Supplementary Results

1.1. Hemispheres considered together
     1.1.1. Anxiety

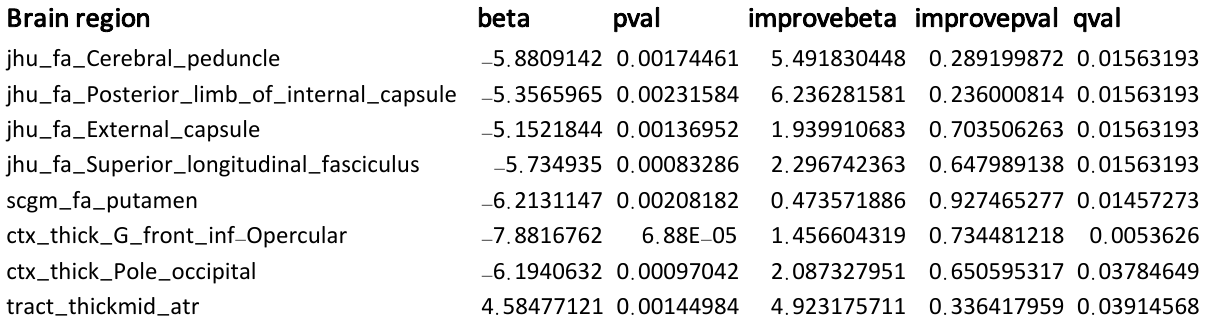

**Table S2**: List of all the brain regions significantly associated with anxiety symptoms

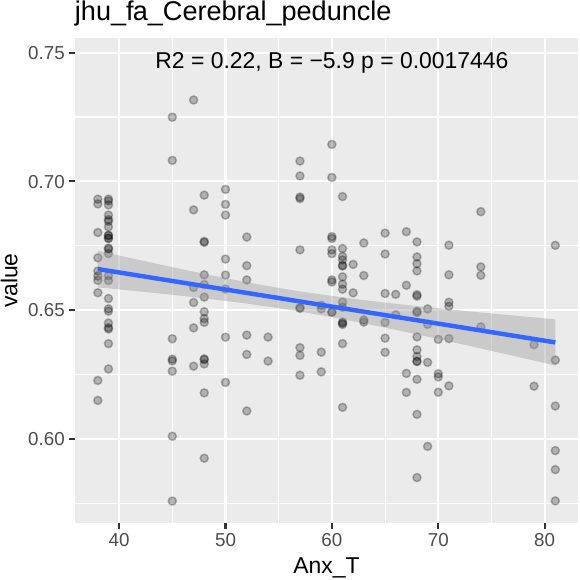

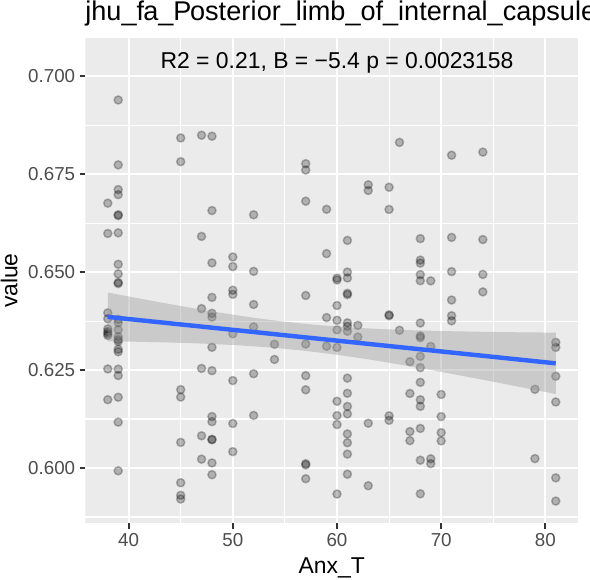

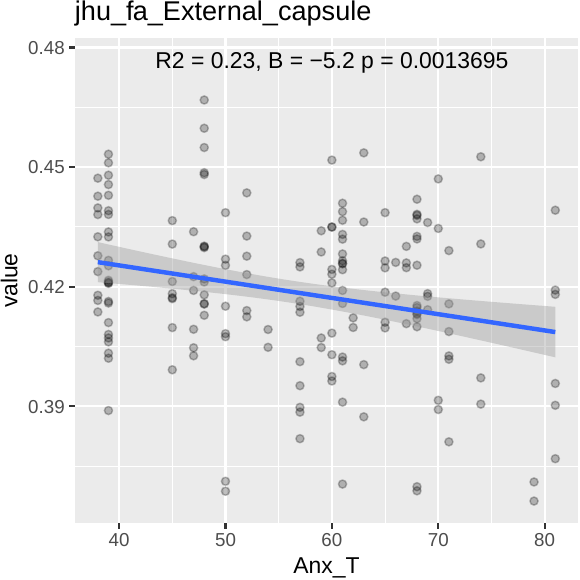

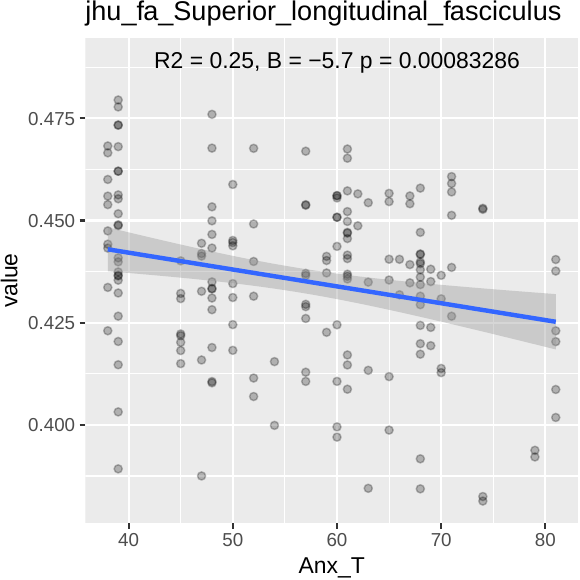

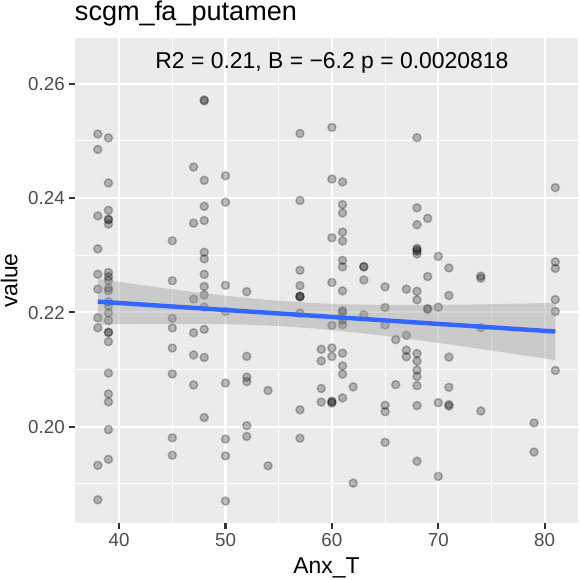

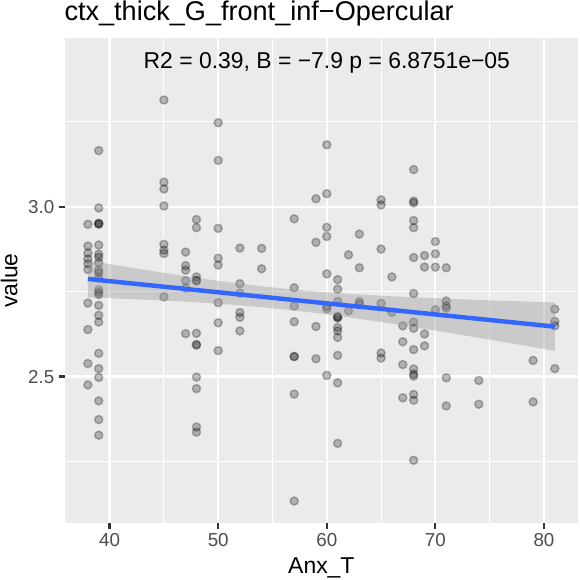

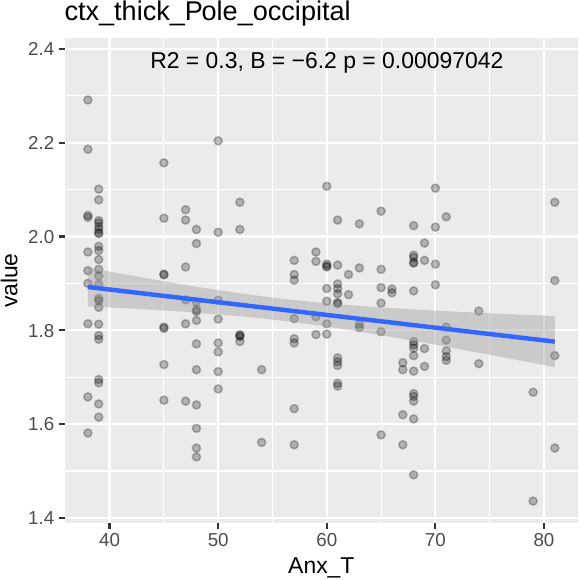

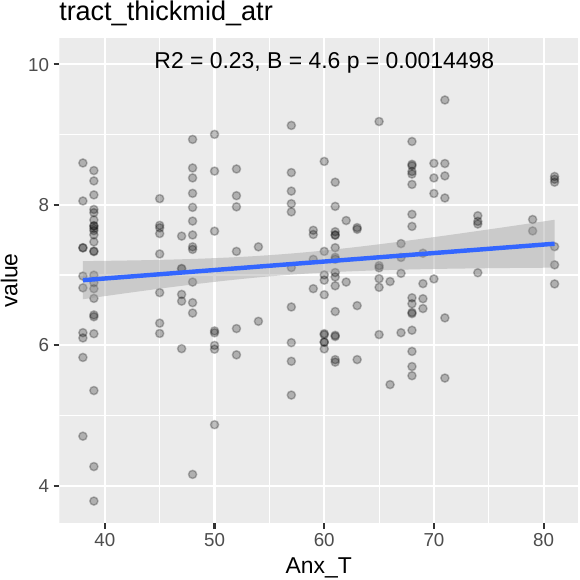

**Figure S1**: Scatterplots indicating brain regions significantly associated to anxiety symptoms (here hemispheres are analyzed together)

1.1.2. Depression

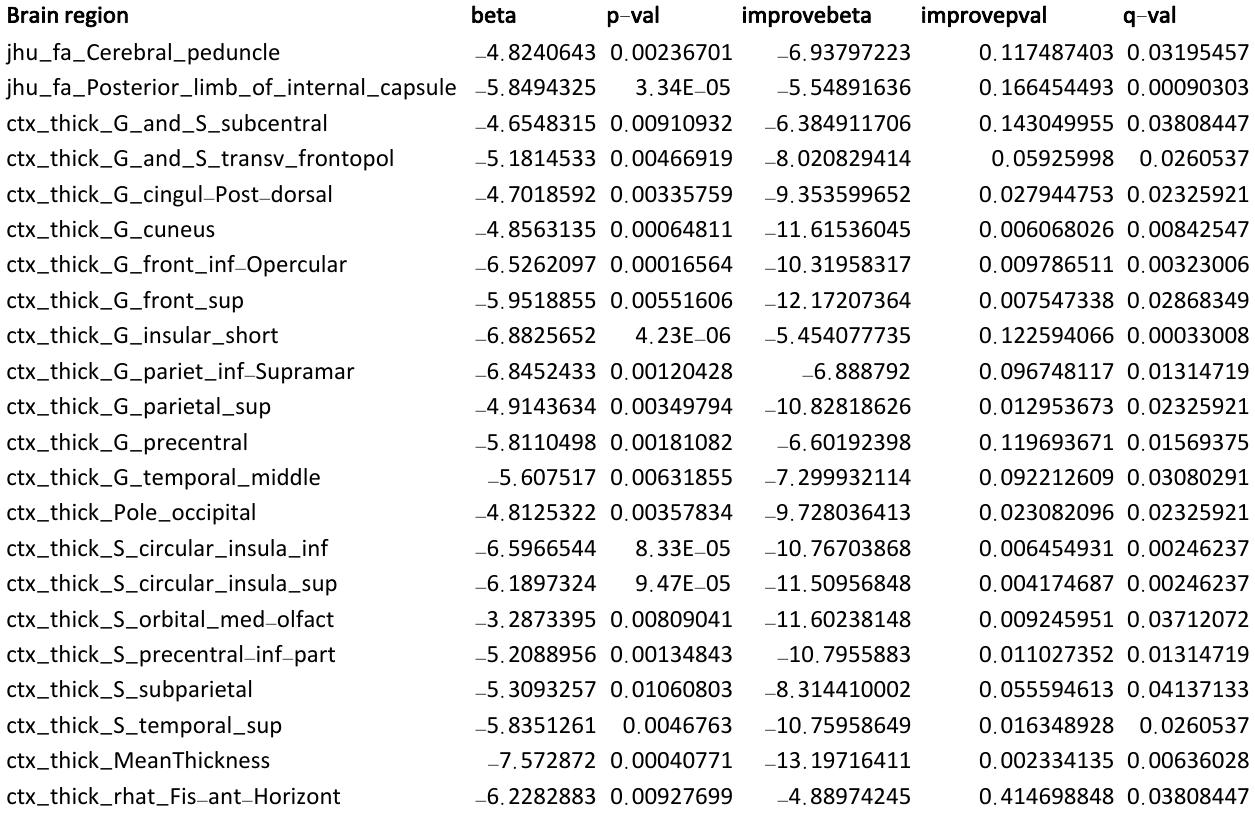

**Table S3**: list of all the brain regions that are significantly associated to depressive symptoms in mTBI patients.

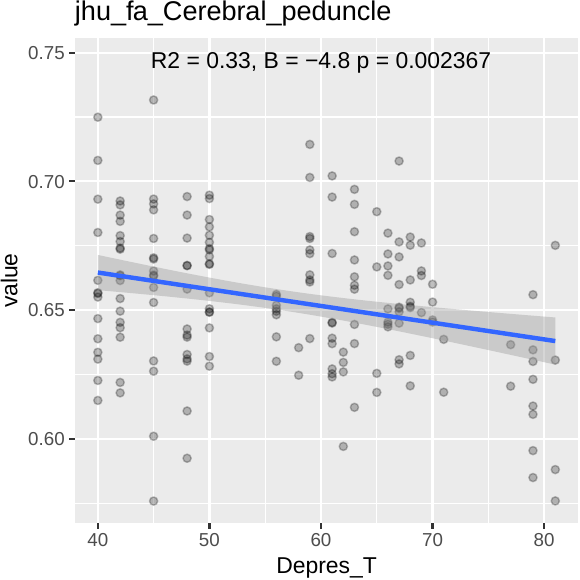

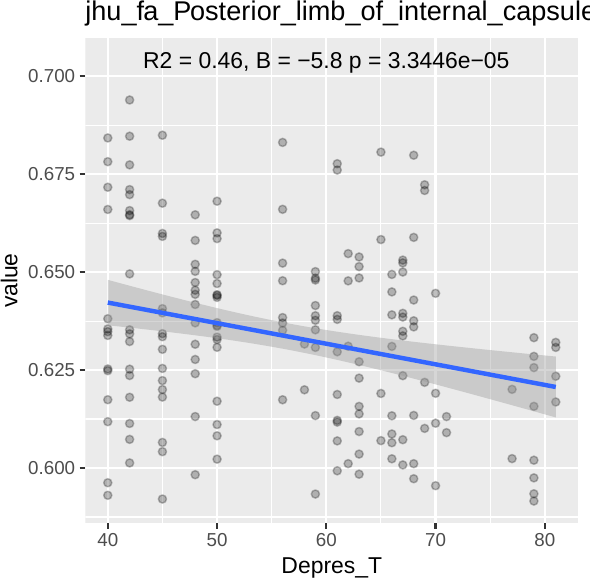

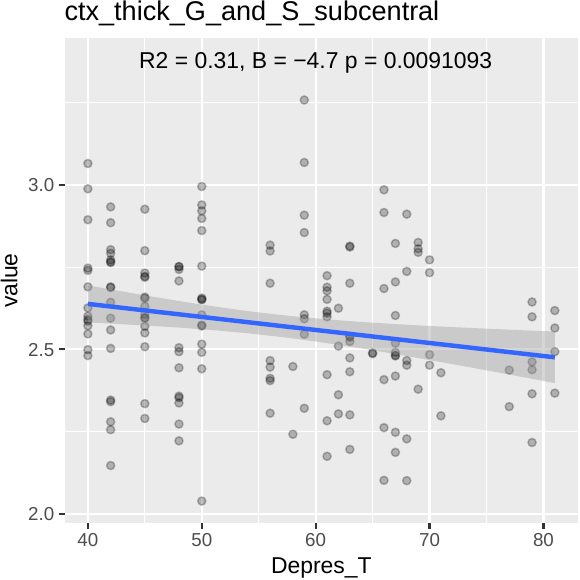

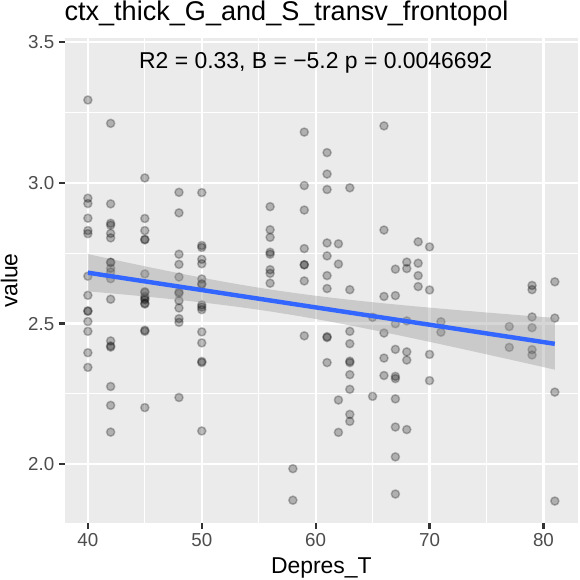

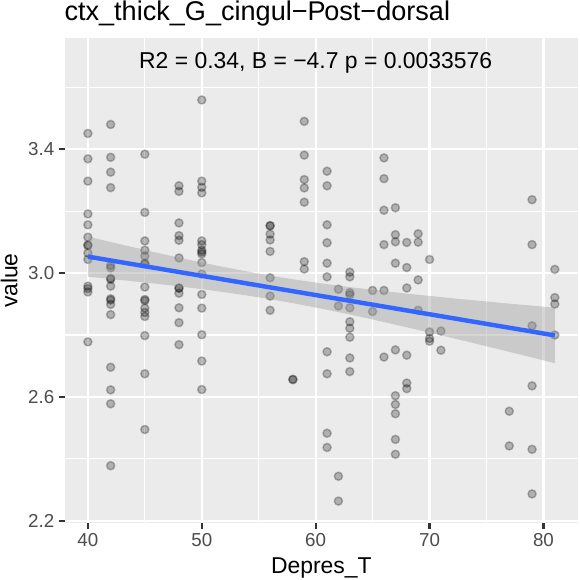

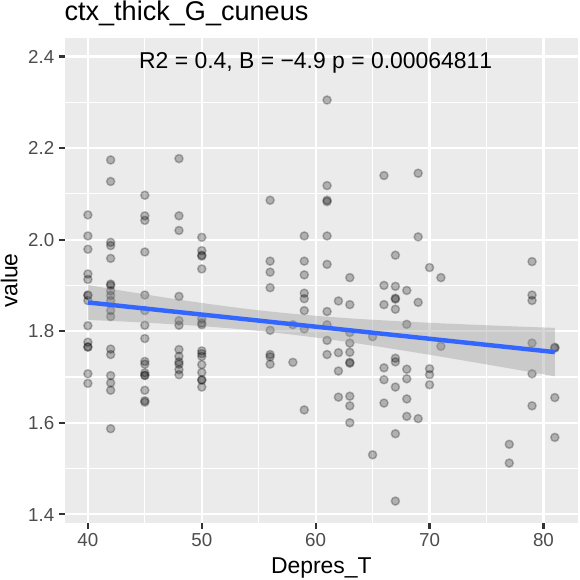

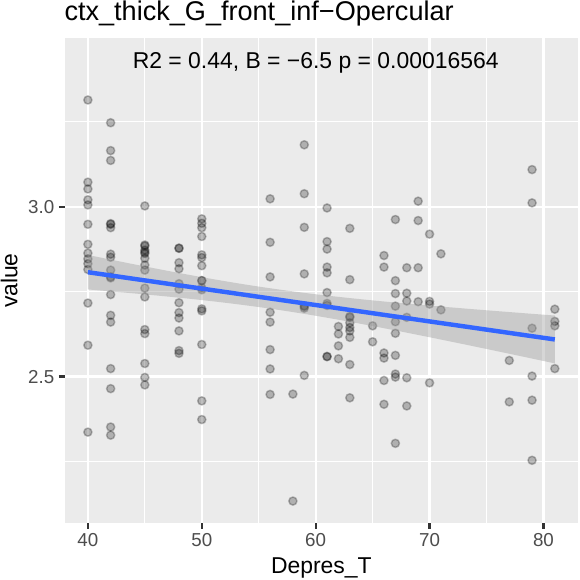

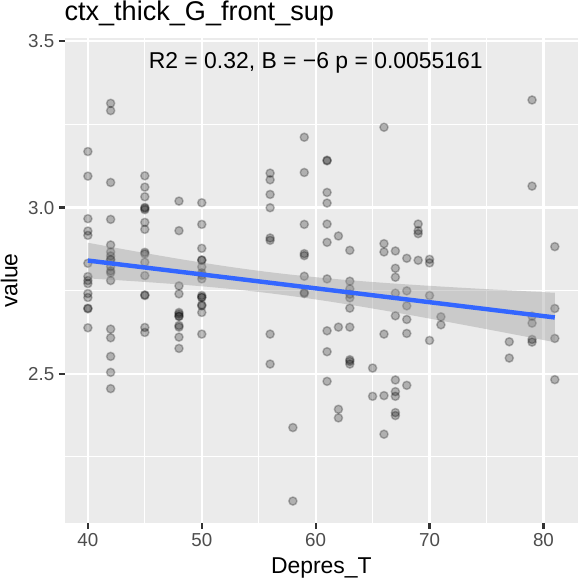

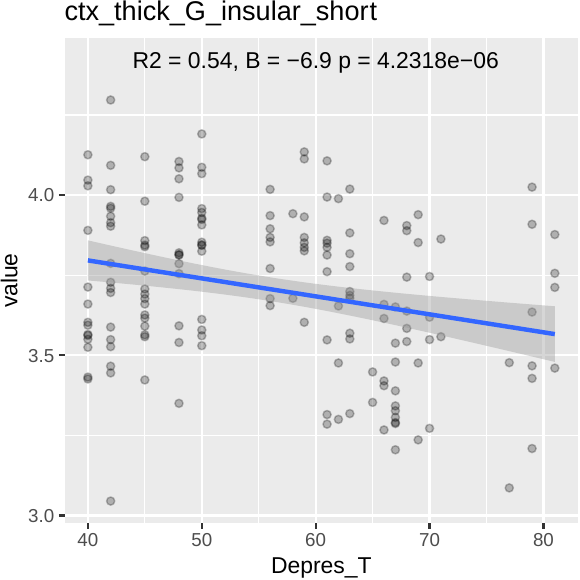

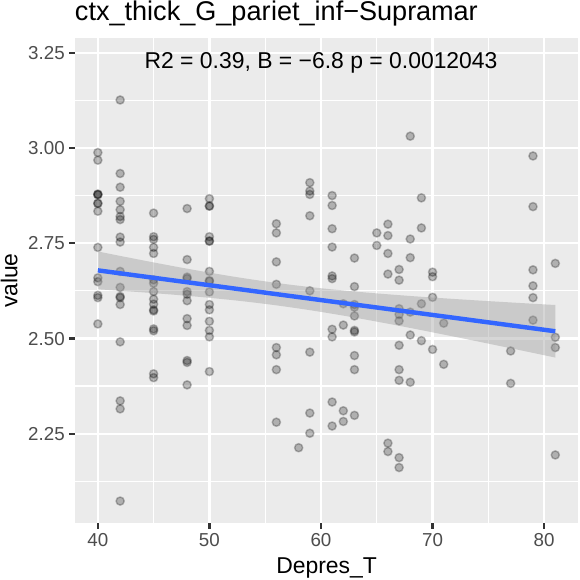

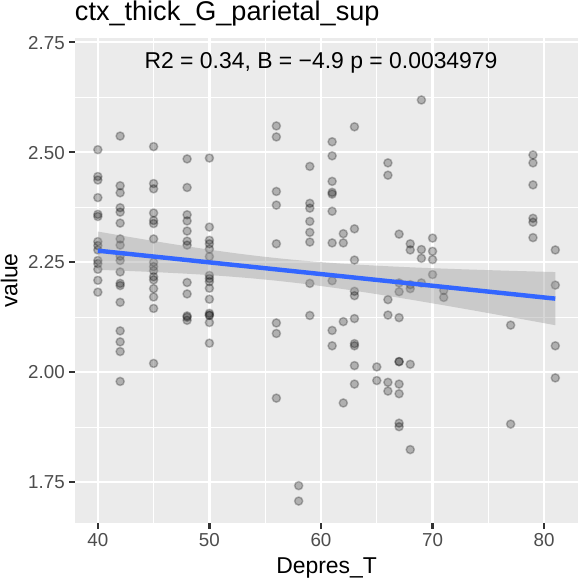

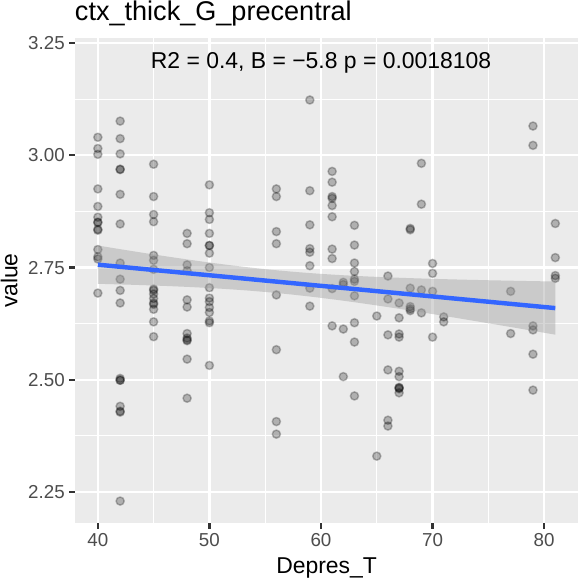

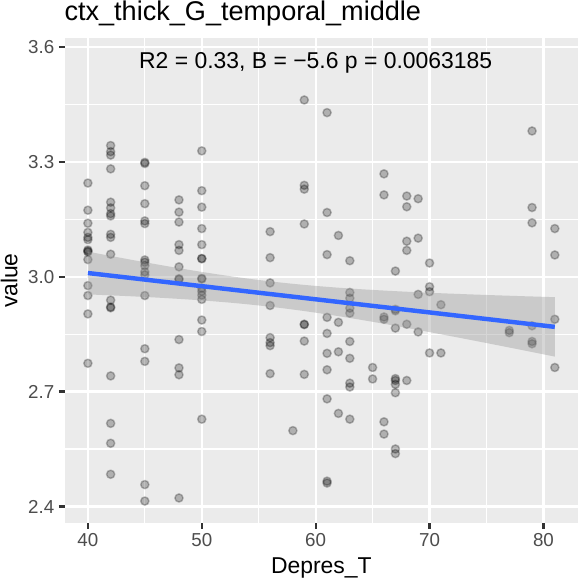

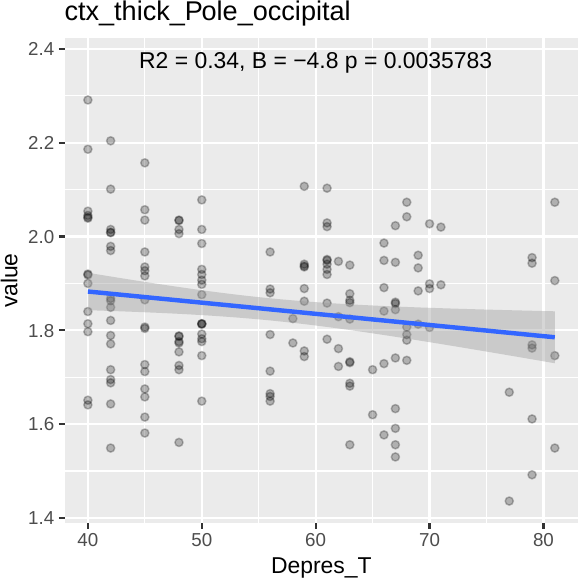

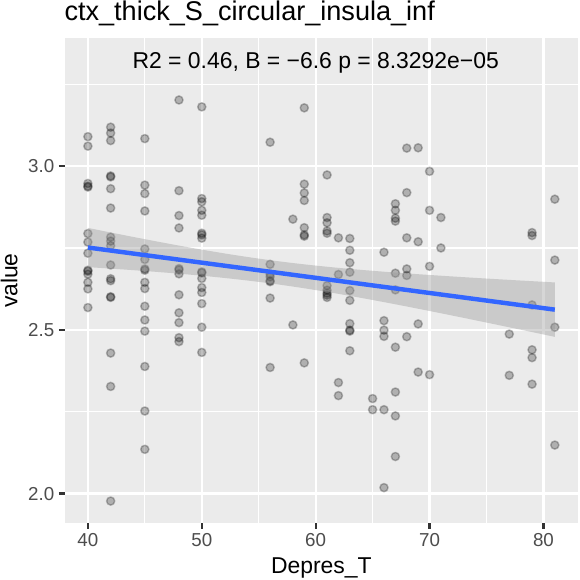

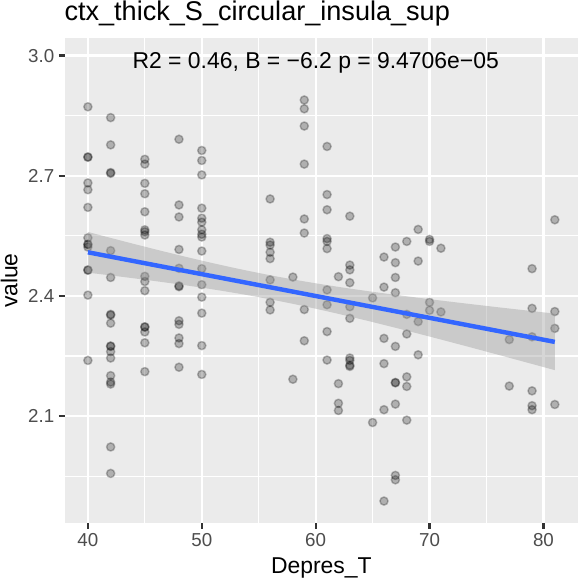

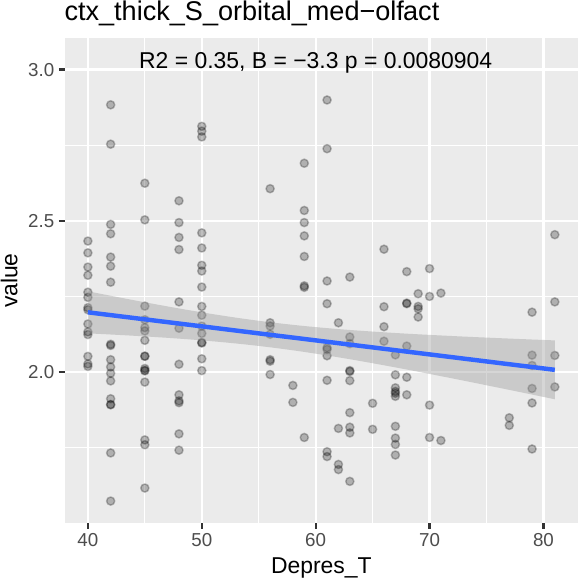

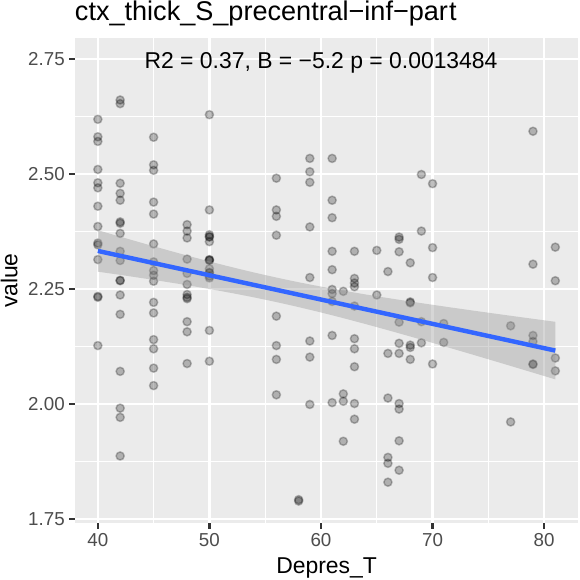

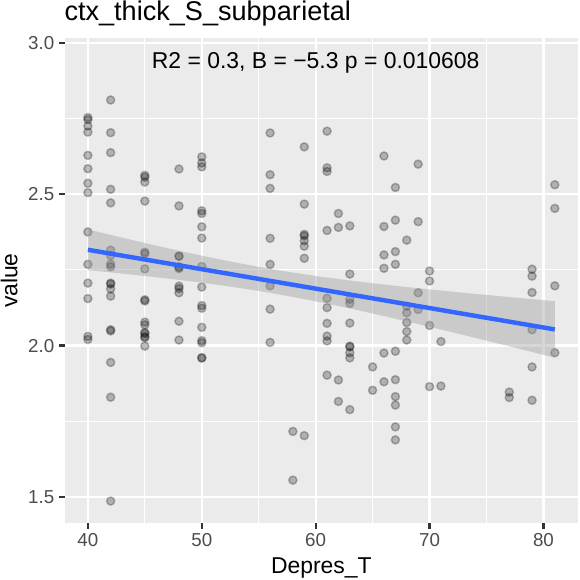

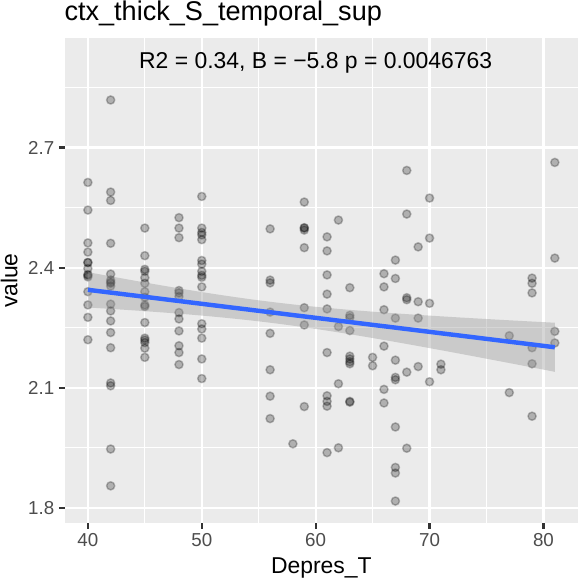

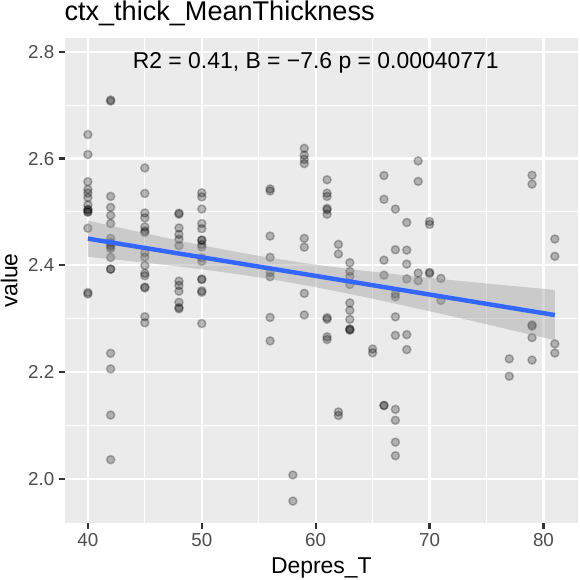

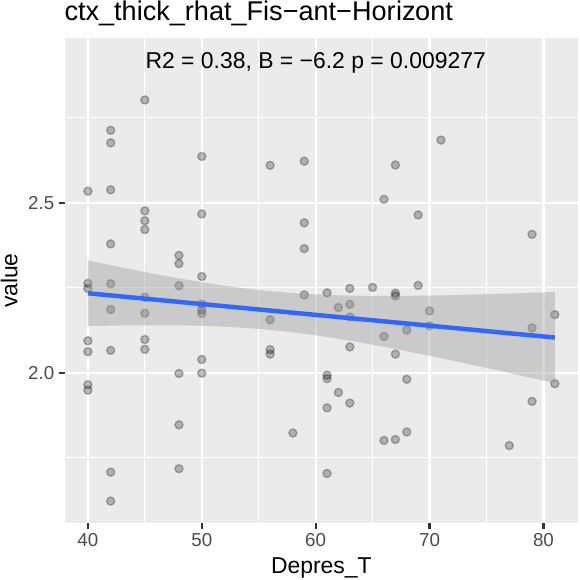

**Figure S2**: Scatterplots indicating the significant relationships between depression and brain regions (here hemispheres are analyzed together)

1.1.3. Somatization

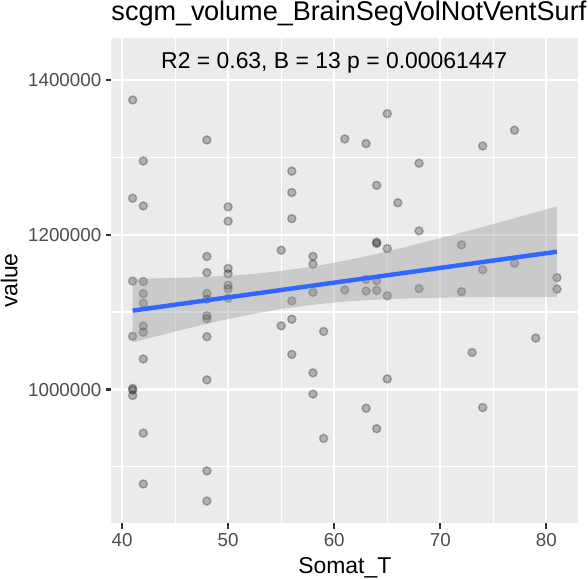

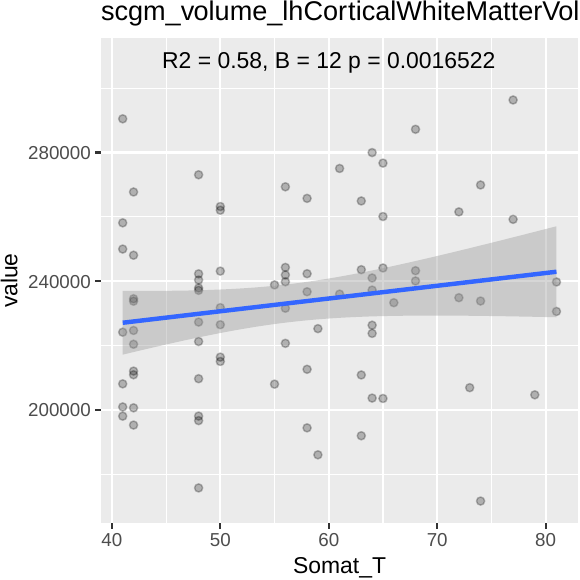

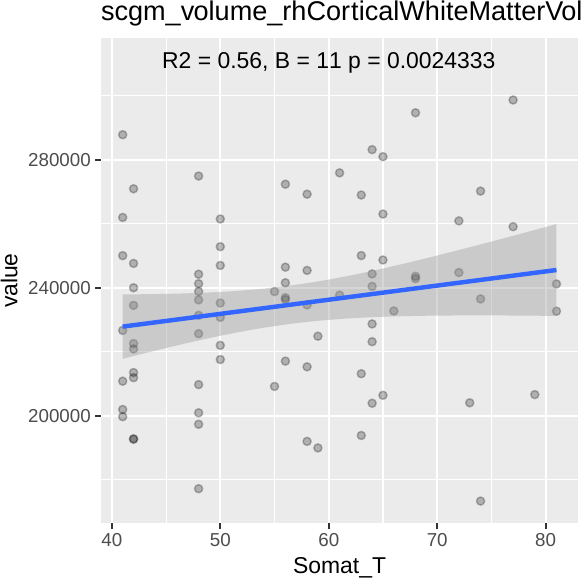

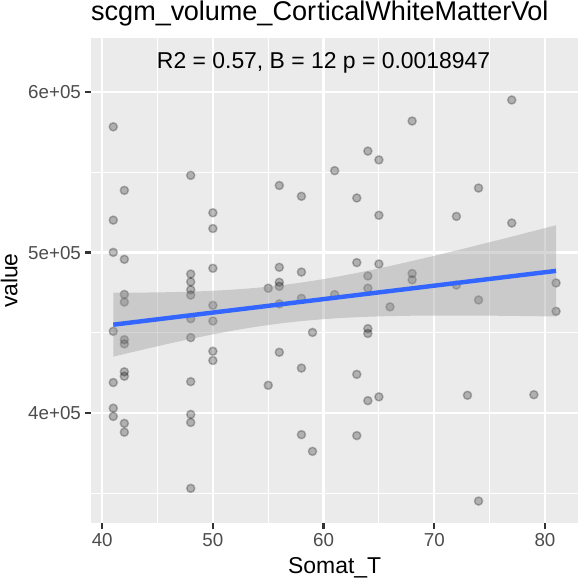

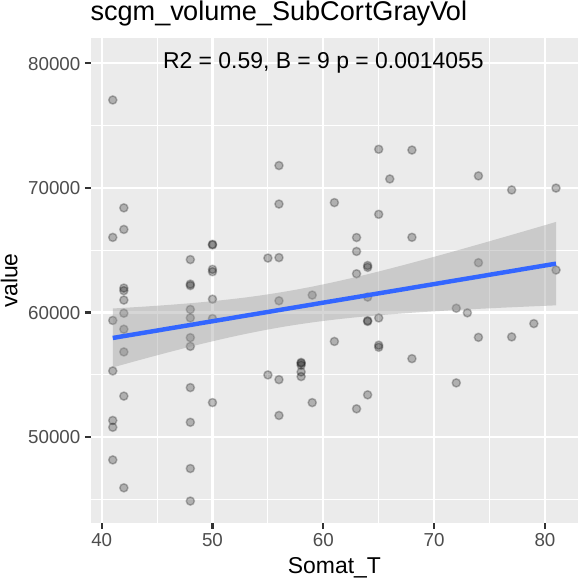

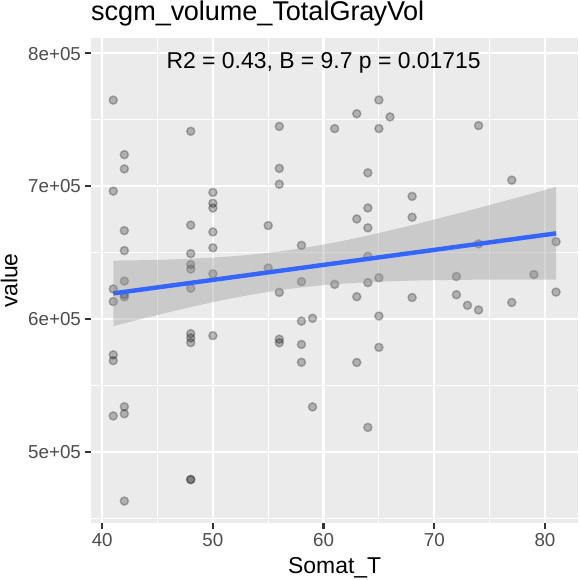

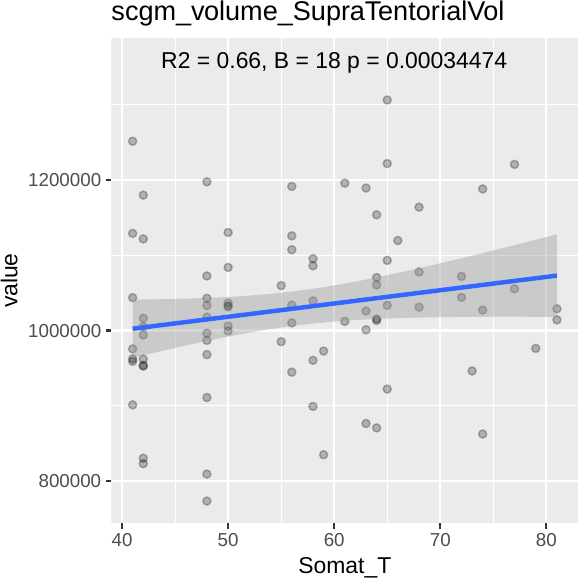

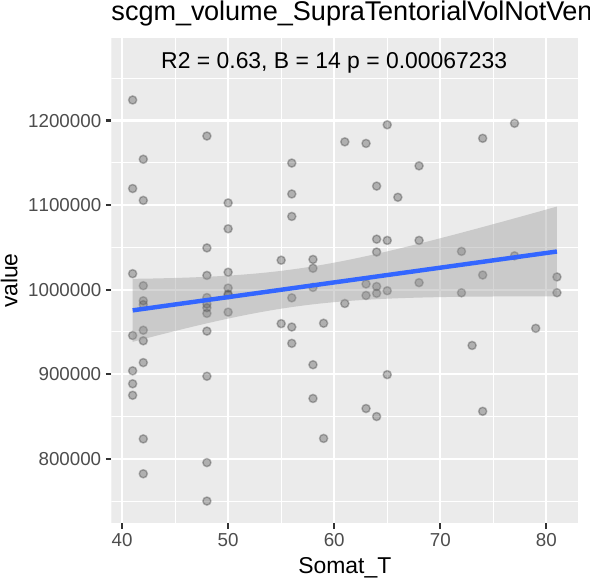

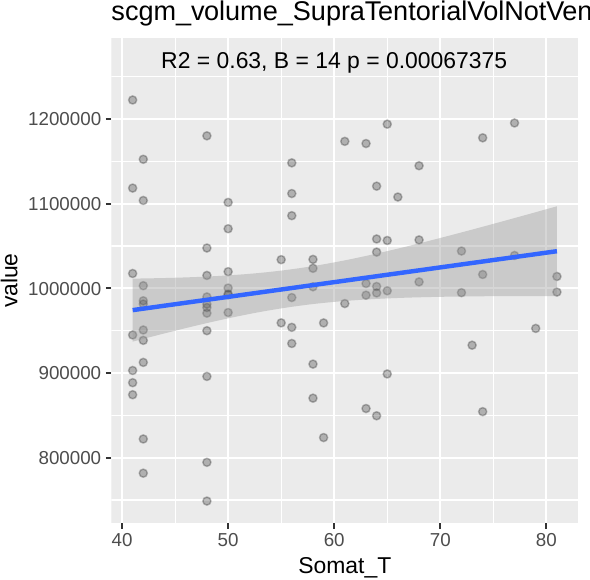

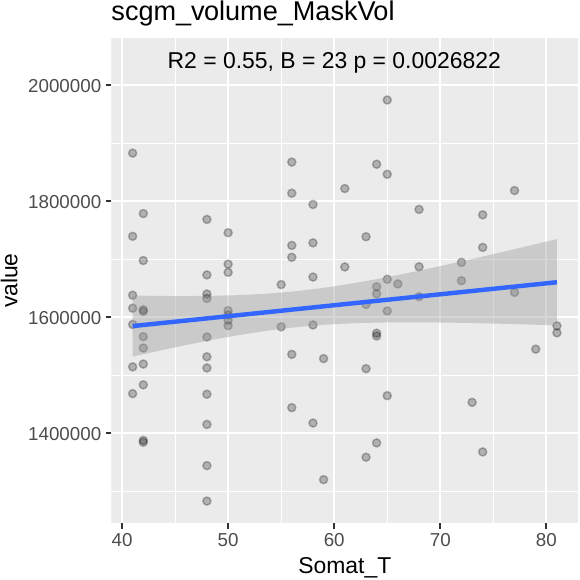

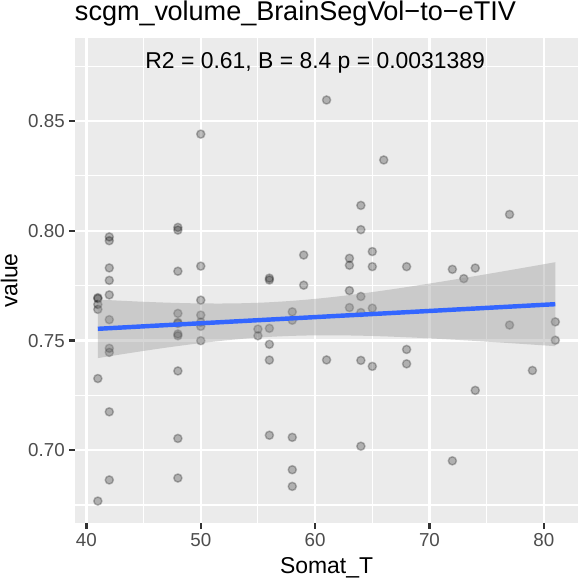

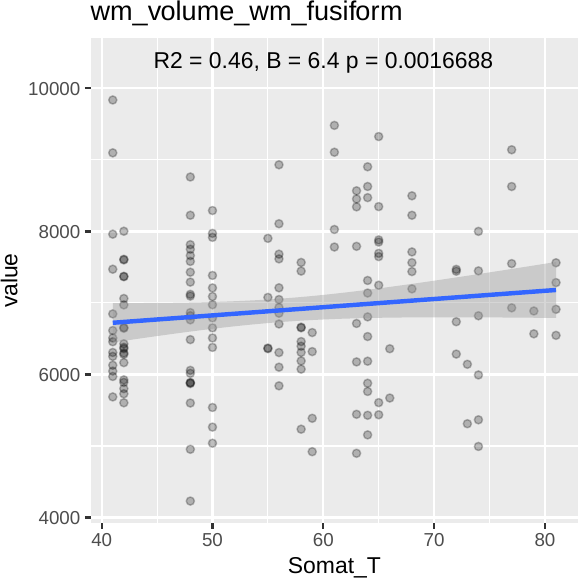

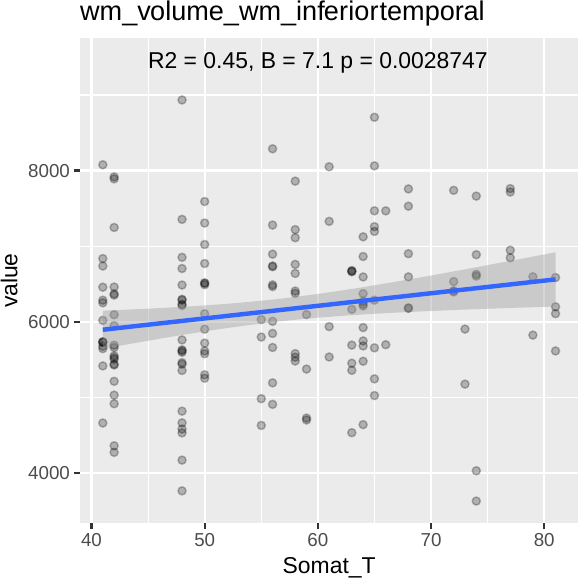

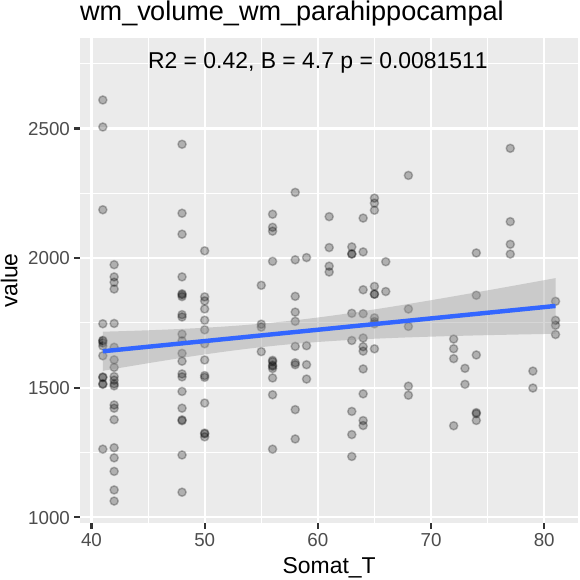

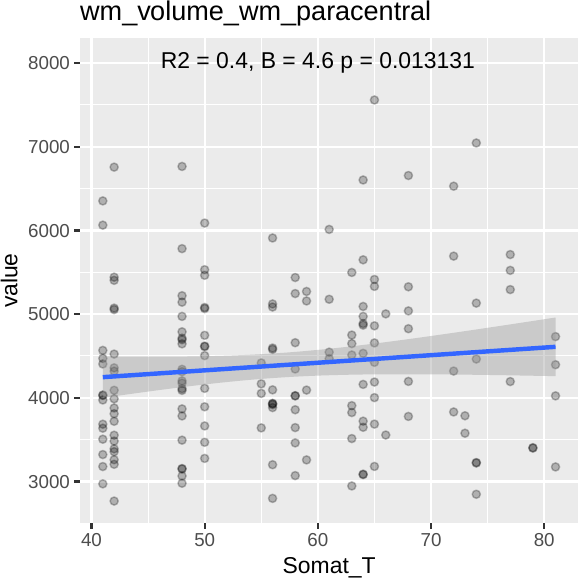

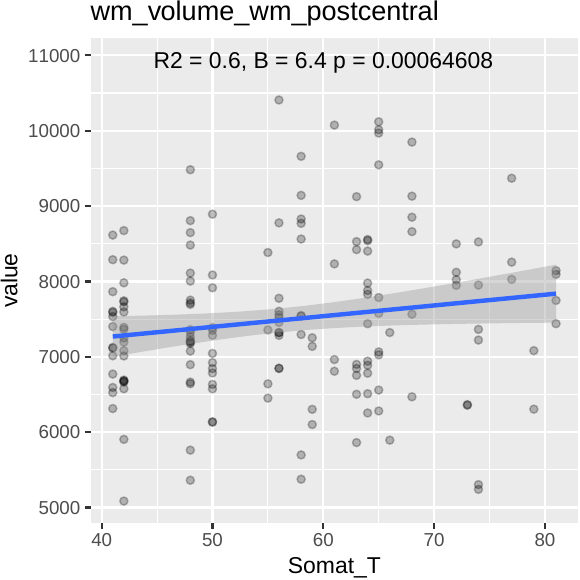

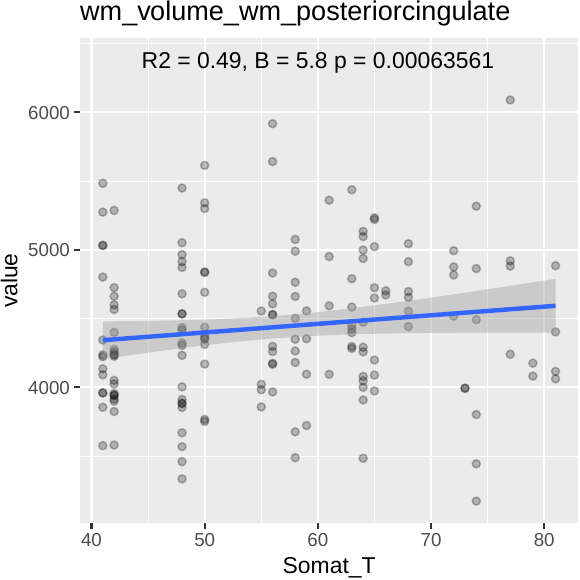

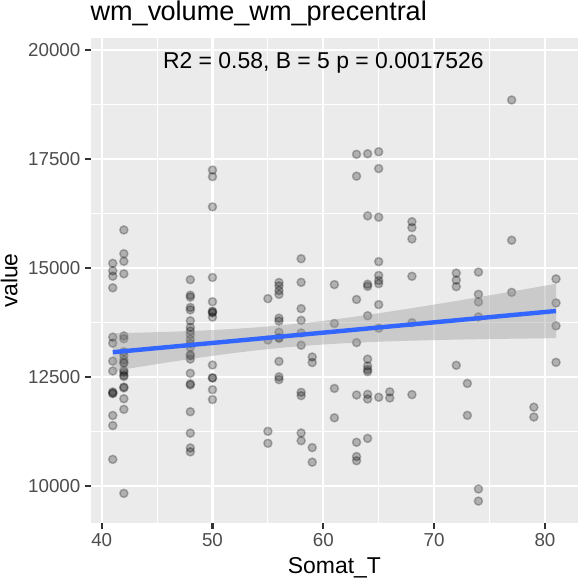

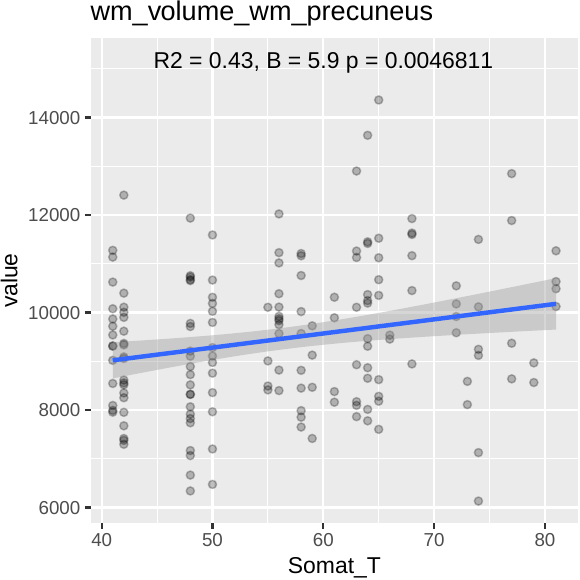

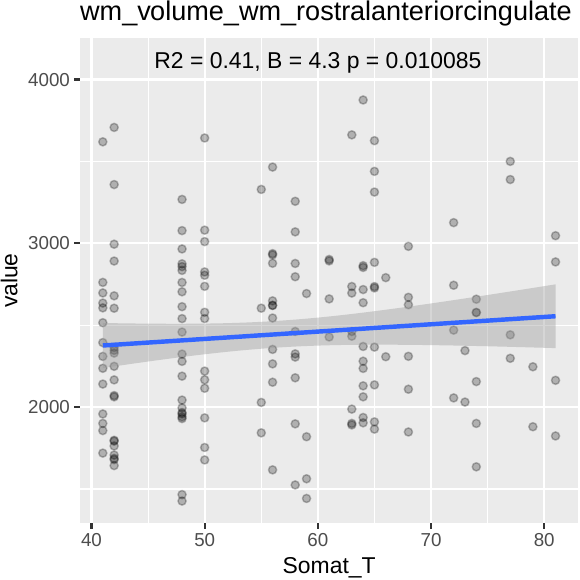

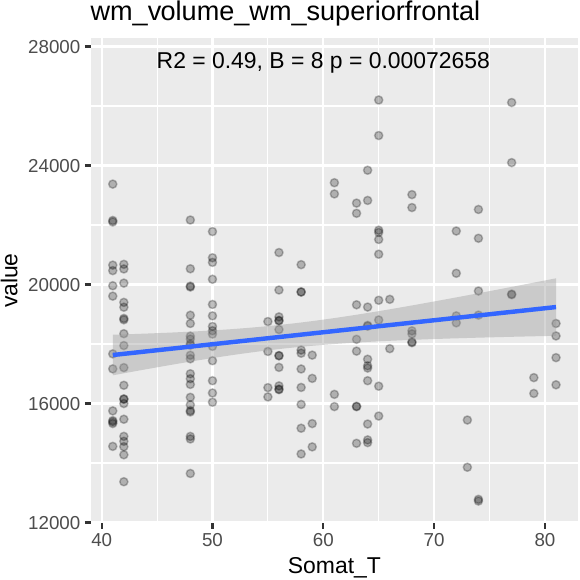

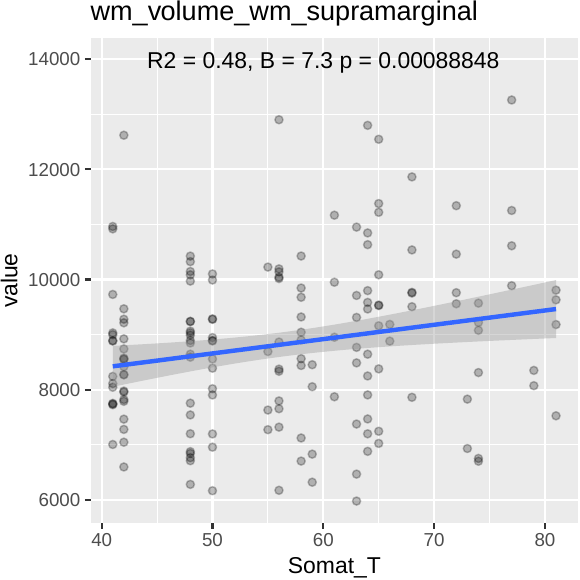

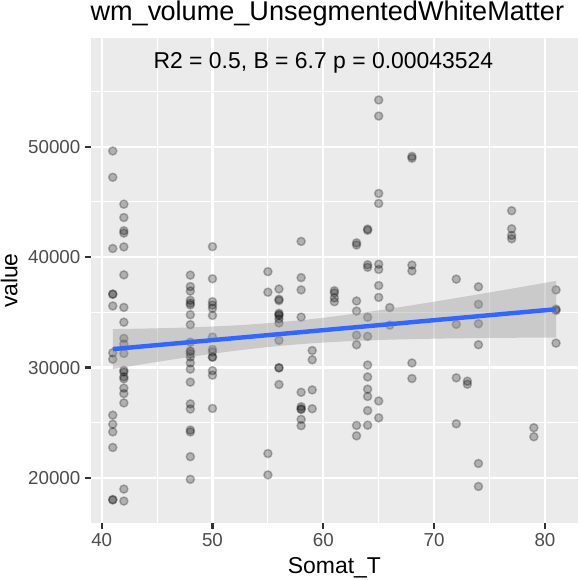

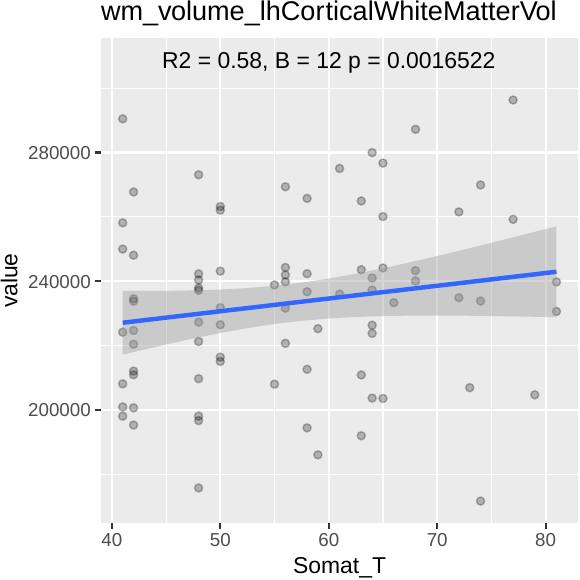

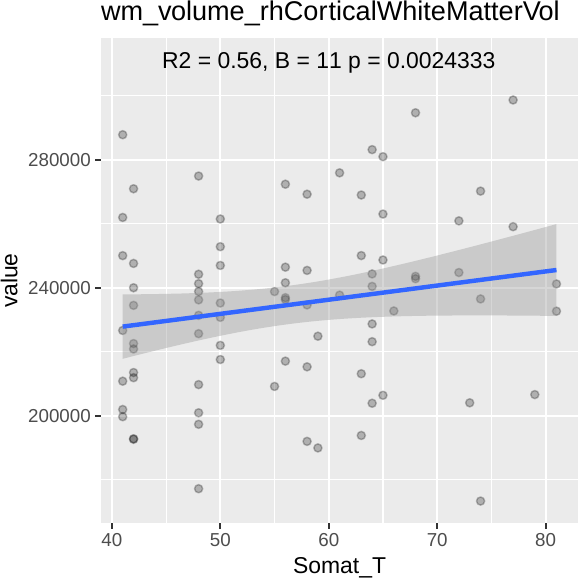

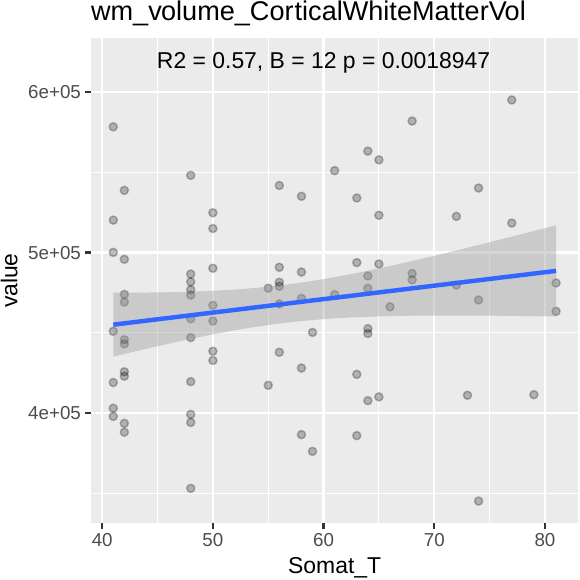

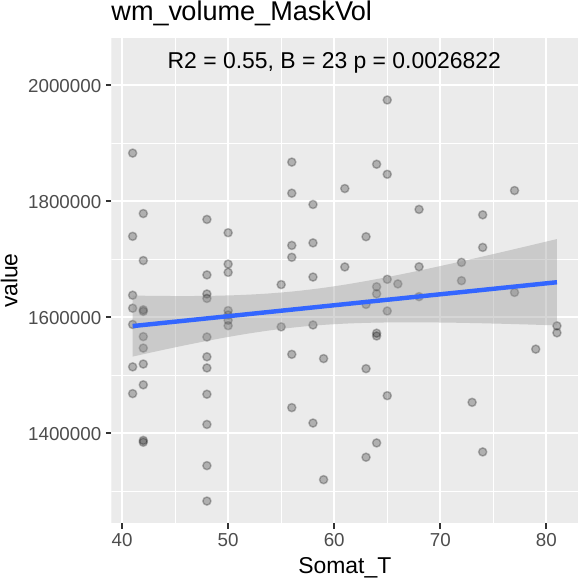

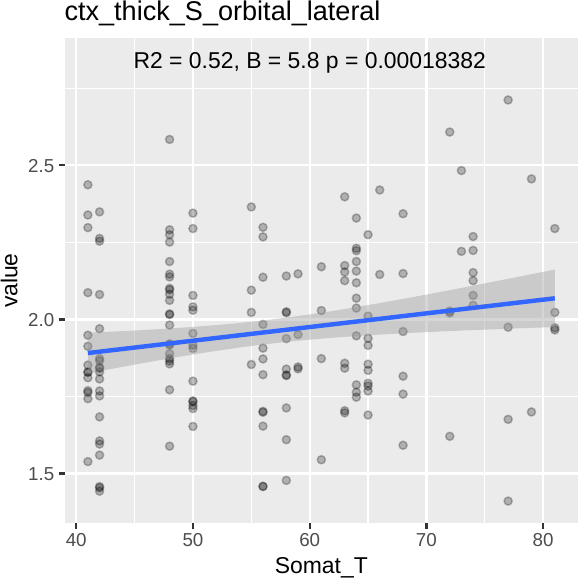

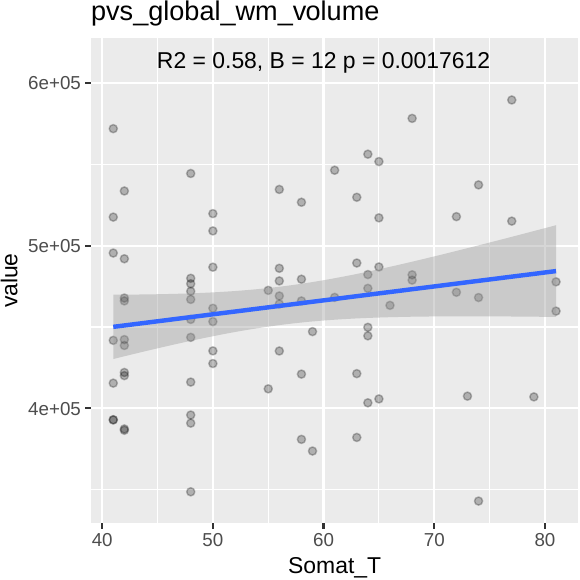

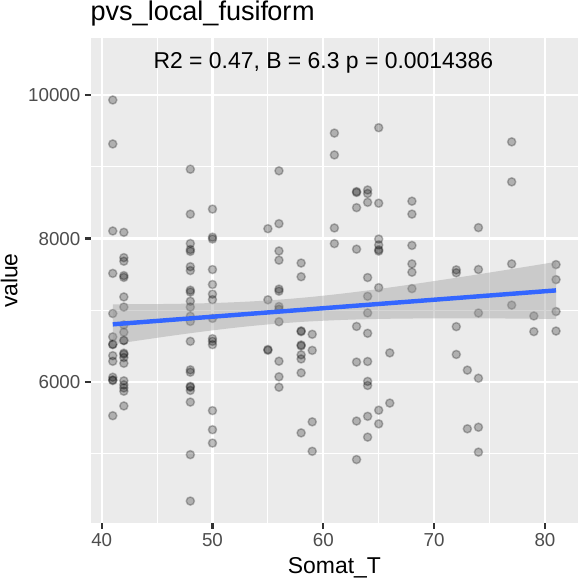

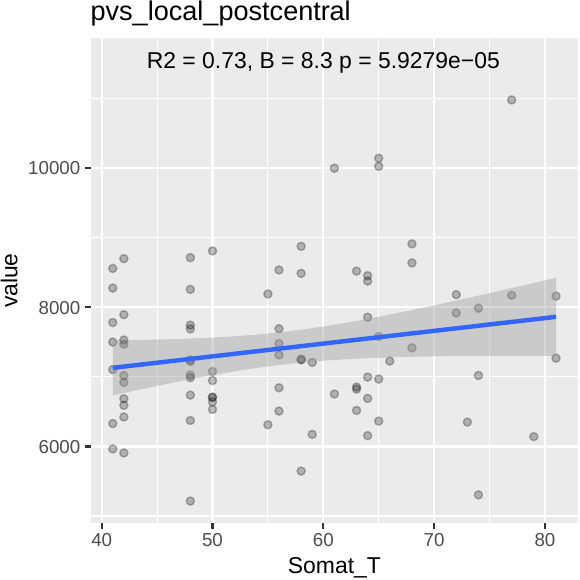

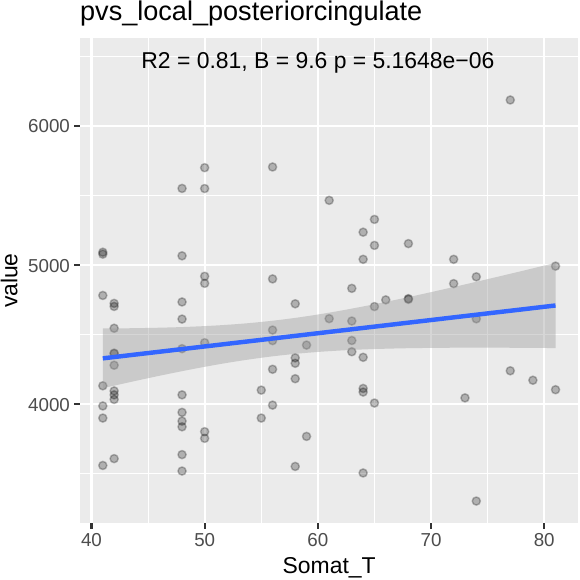

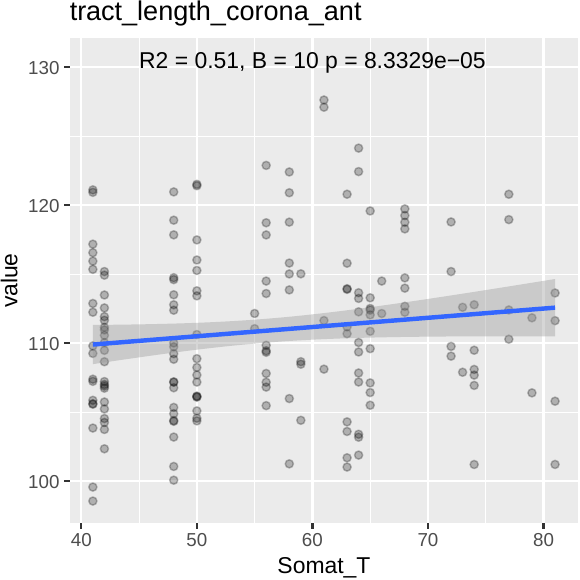

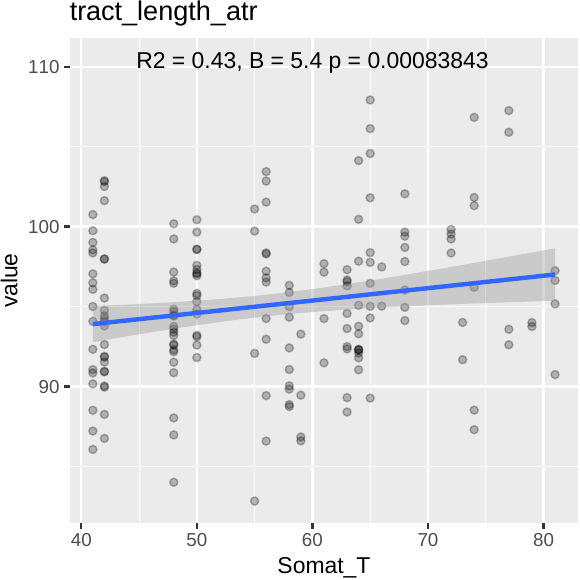

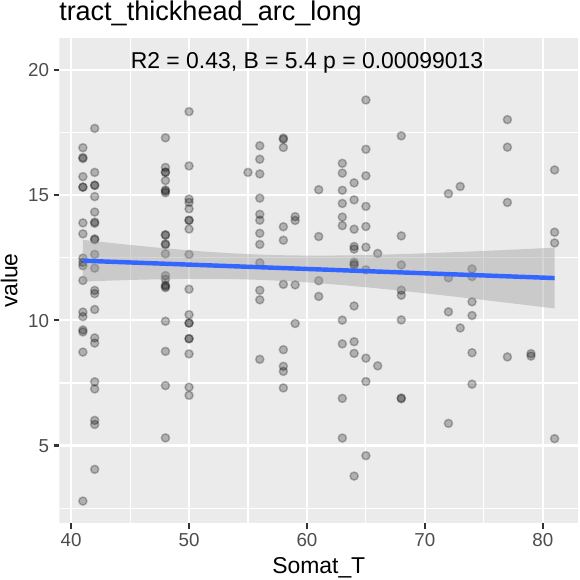

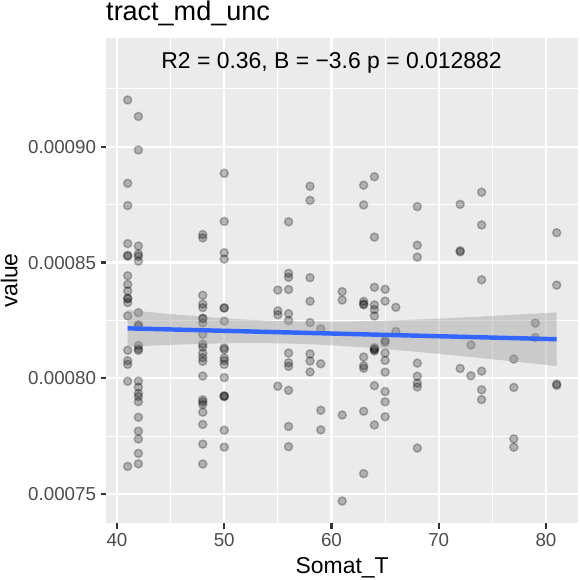

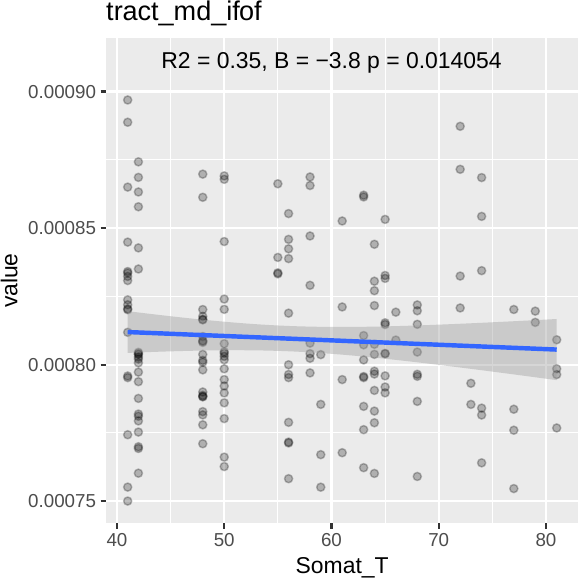

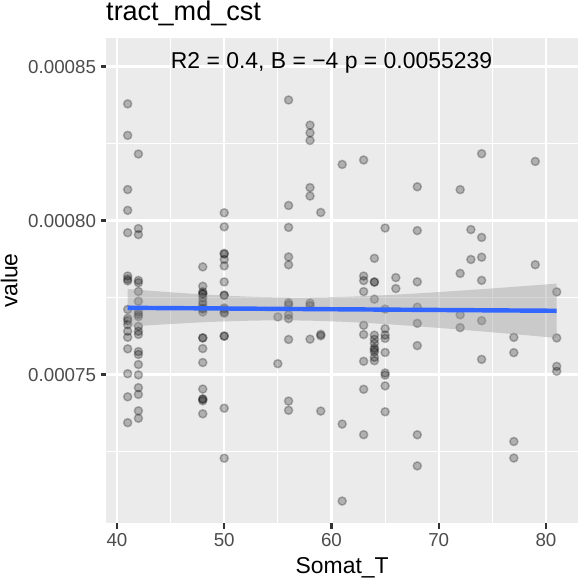

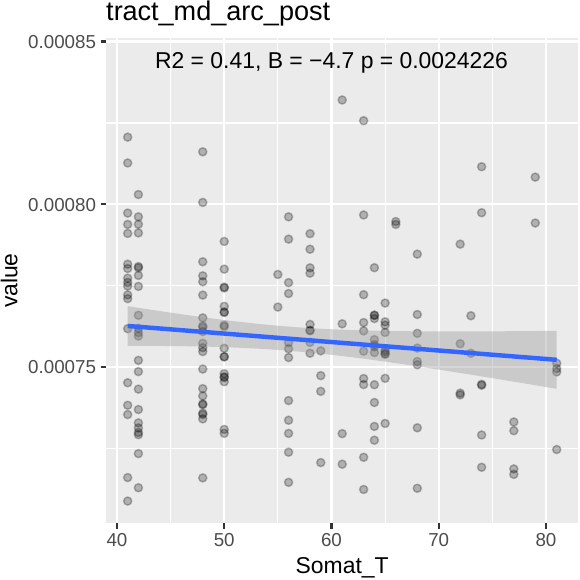

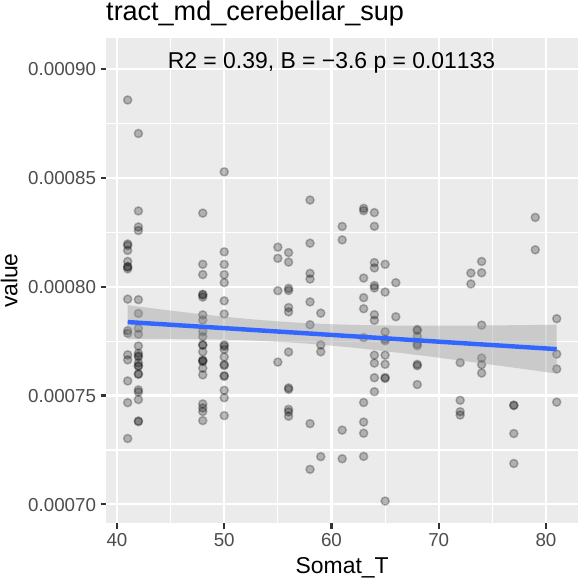

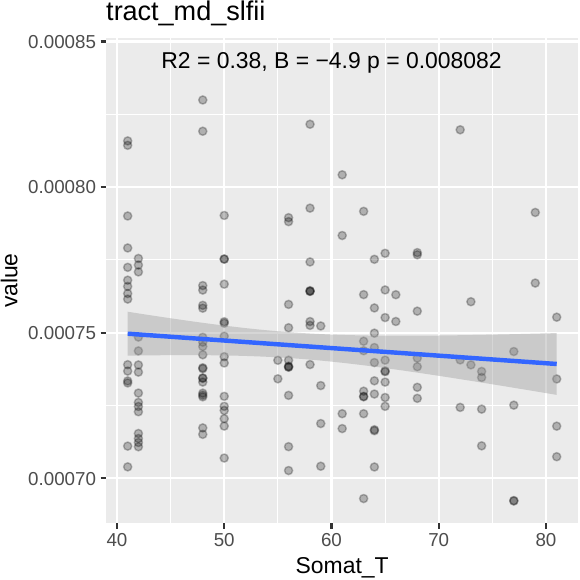

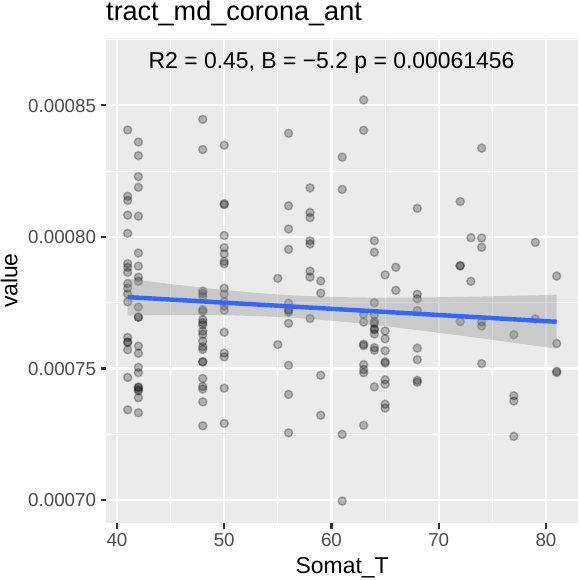

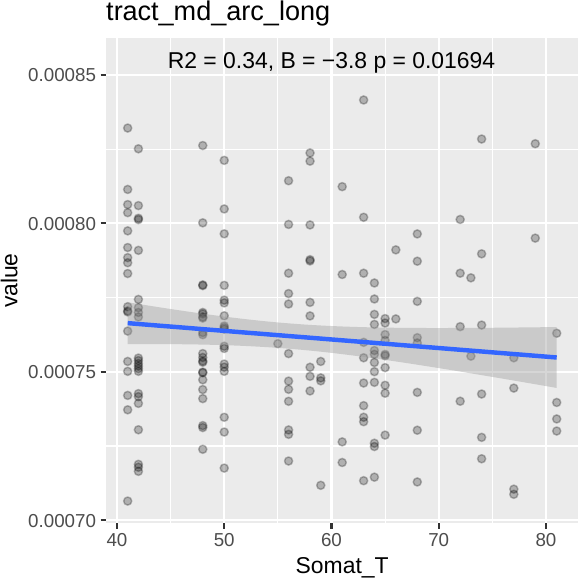

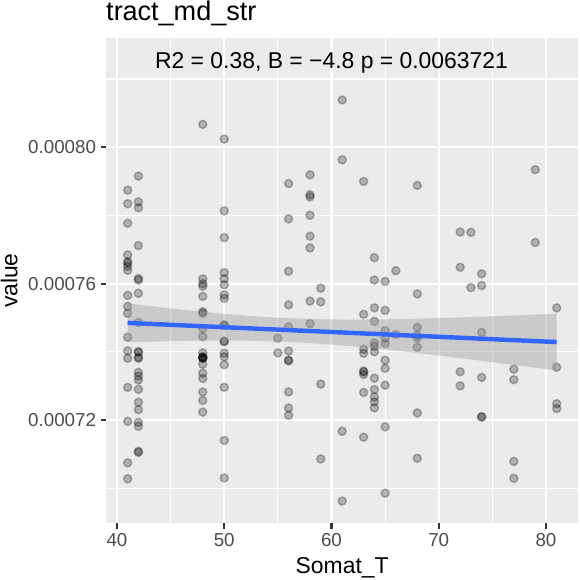

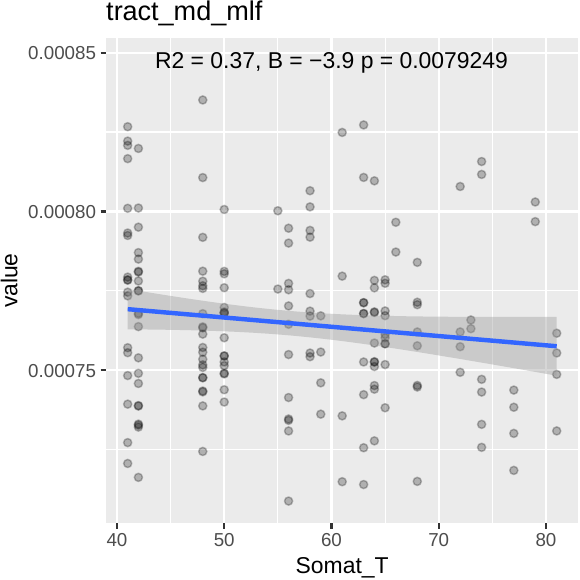

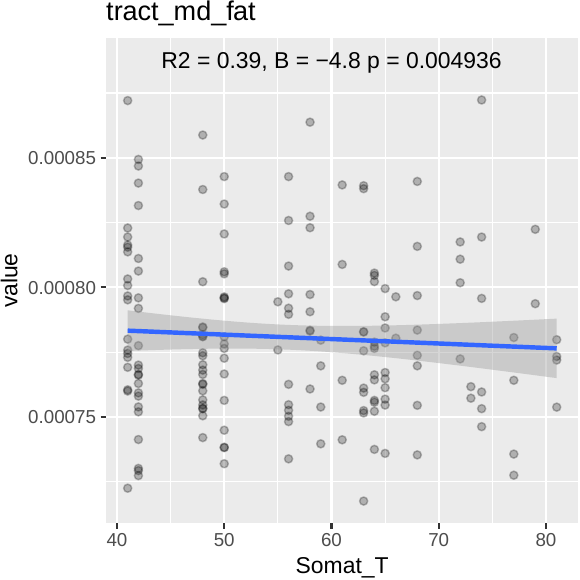

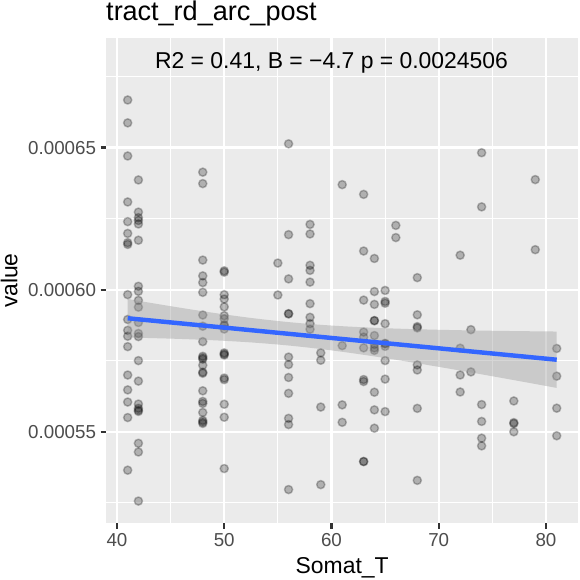

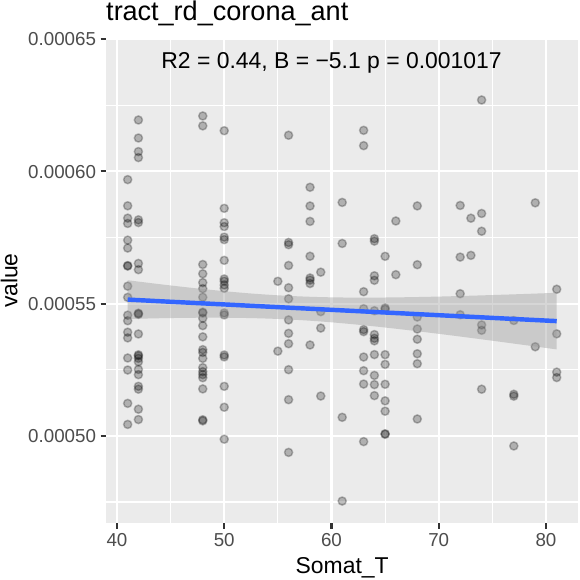

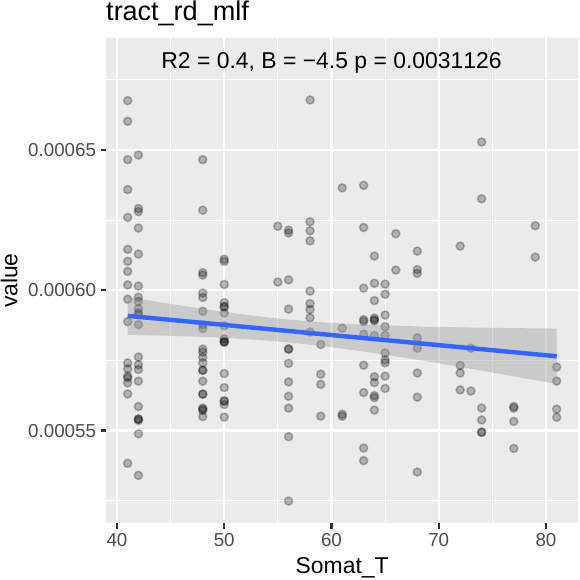

**Figure S3**: Scatterplots indicating statistically significant relationships between somatization symptoms and brain regions (here hemispheres are analyzed together)

1.2. Hemispheres separated

1.2.1. Depression and anxiety lateralization

For depression and anxiety symptoms, there were no statistically significant associations when considering the two hemispheres together, after FDR correction for multiple comparisons (p>0.05).

1.2.2. Somatization

Taking into account hemispheric lateralization, somatization symptoms were positively associated to the volume of the left thalamus, hippocampus, ventral diencephalon (formed by hypothalamus, mammillary body, subthalamic nuclei, substantia nigra, red nucleus, lateral geniculate nucleus, and medial geniculate nucleus). In the right hemisphere, significant positive relationships were seen with the volume of hypothalamus, amygdala, the white matter of posterior cingulate, and the volume of subcortical structures. Globally, volumes of total white matter and subcortical grey matter were affected by somatization.

For the PVS results, local significant positive associations were found in the left postcentral and right posterior cingulate. Regarding white matter tracts, there was a positive relationship between the thickness of the right arcuate fasciculus and the left superior thalamic radiation.

Below scatterplots of significant relationships are shown.

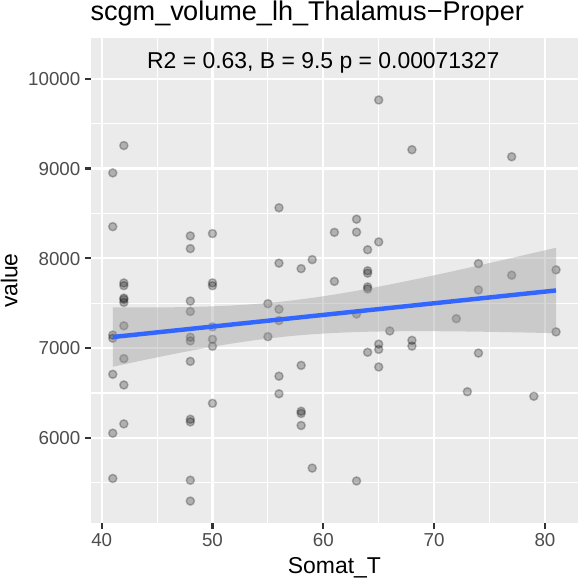

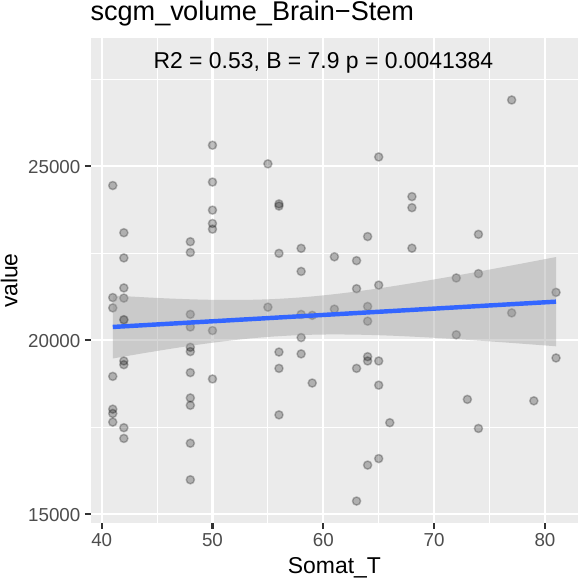

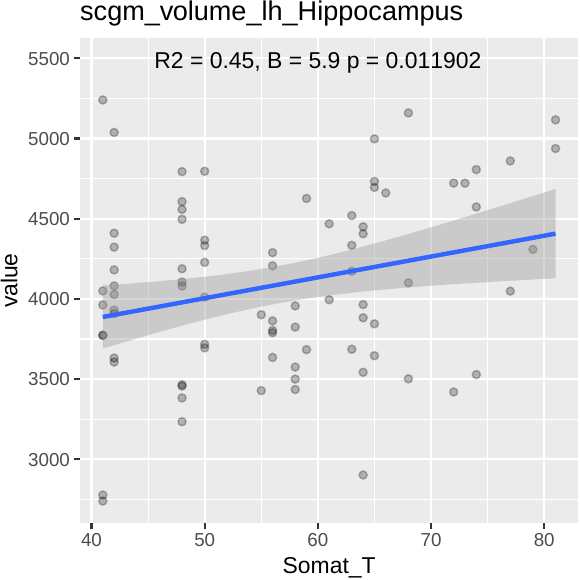

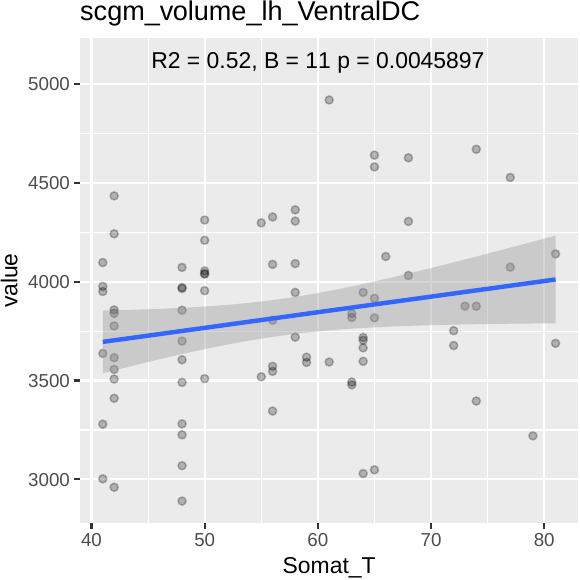

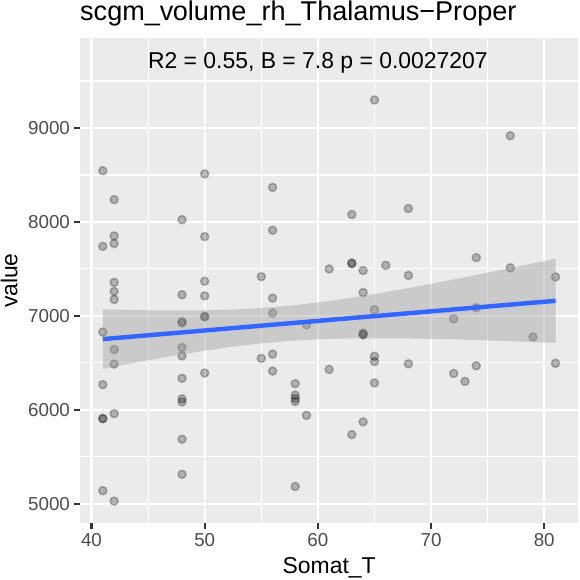

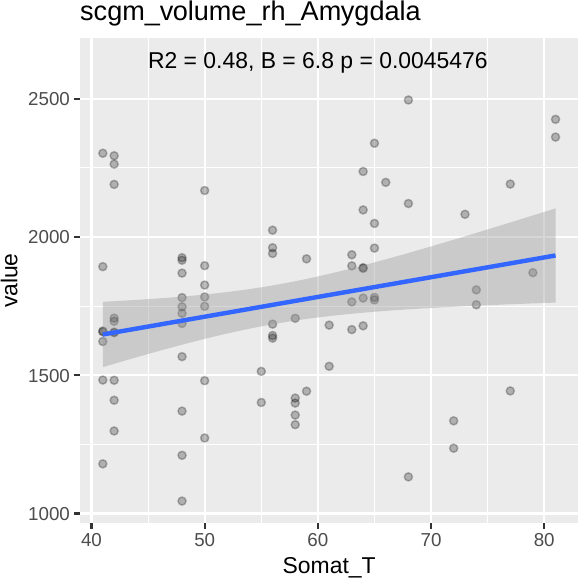

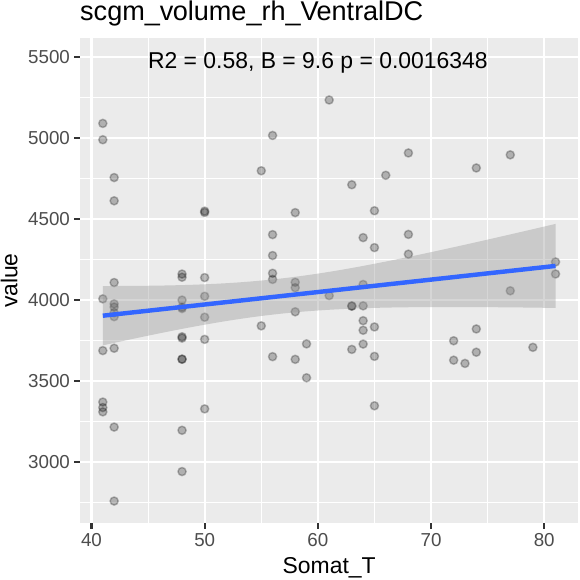

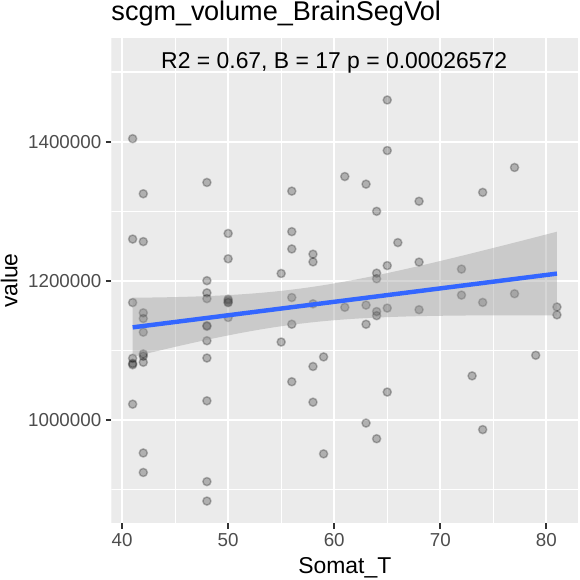

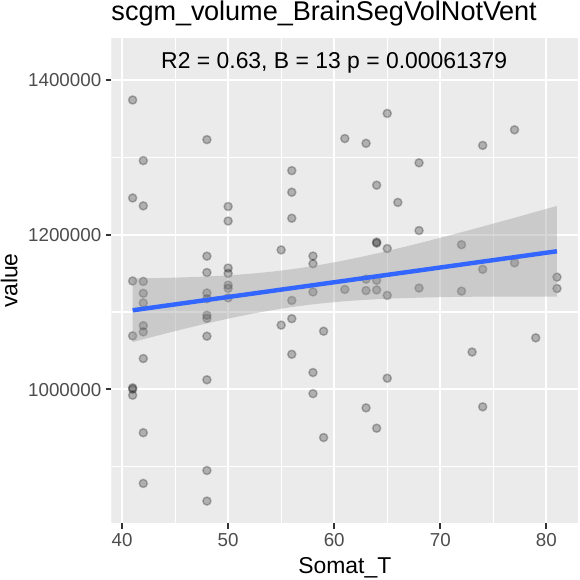

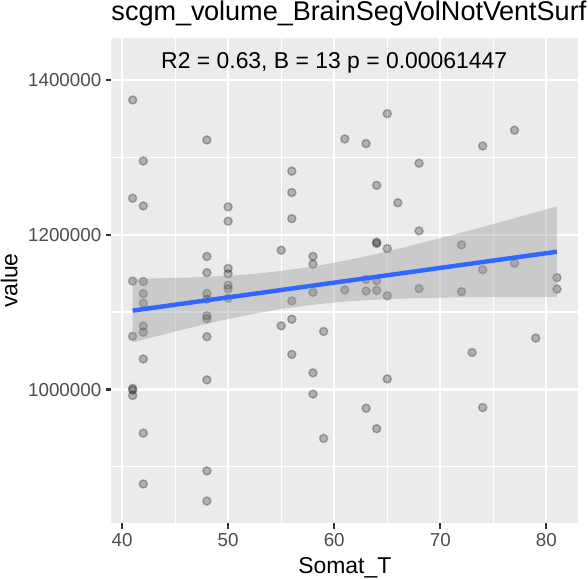

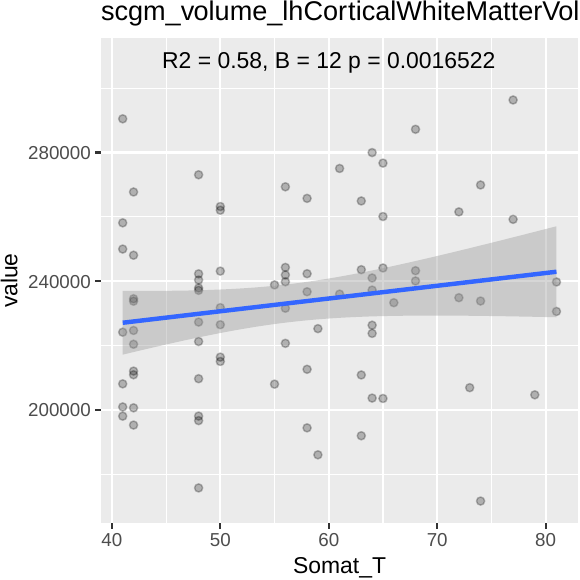

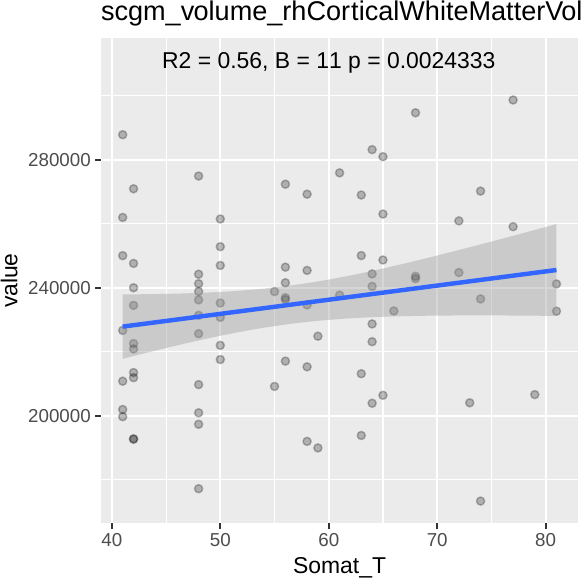

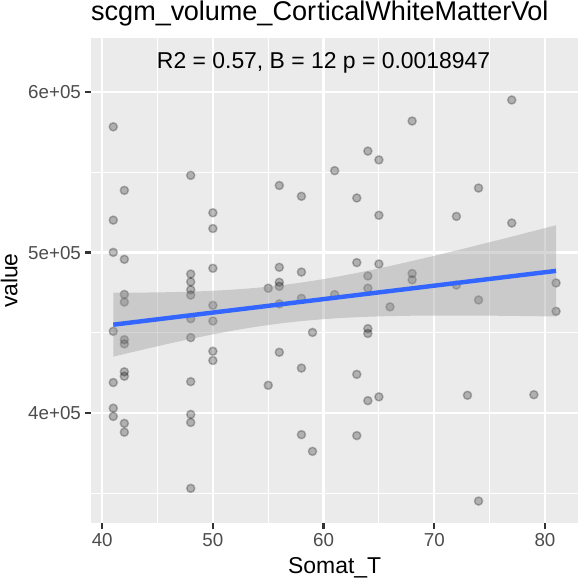

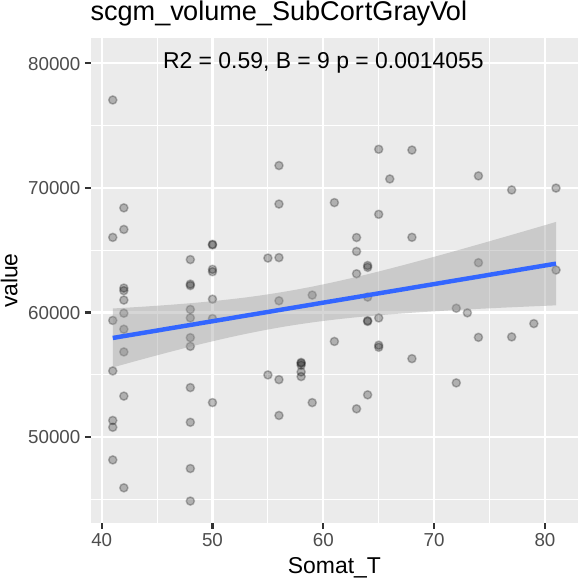

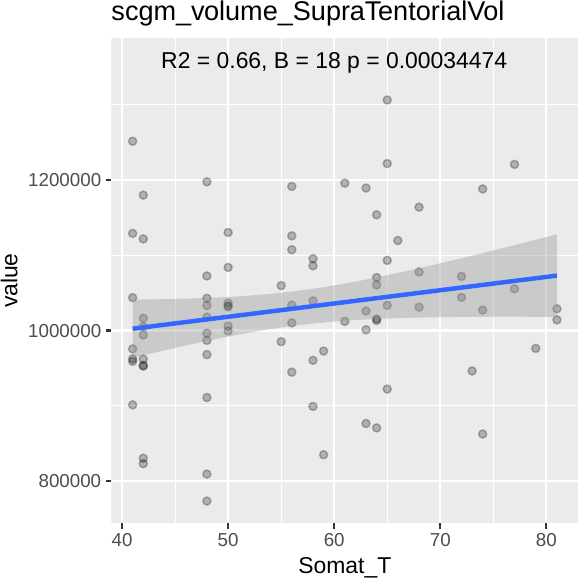

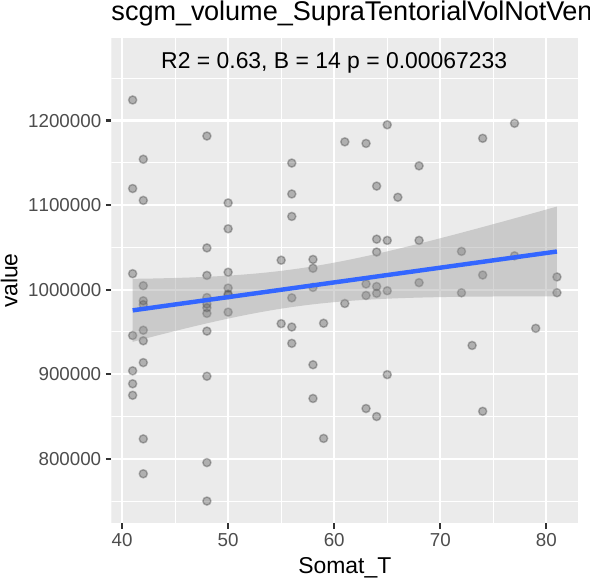

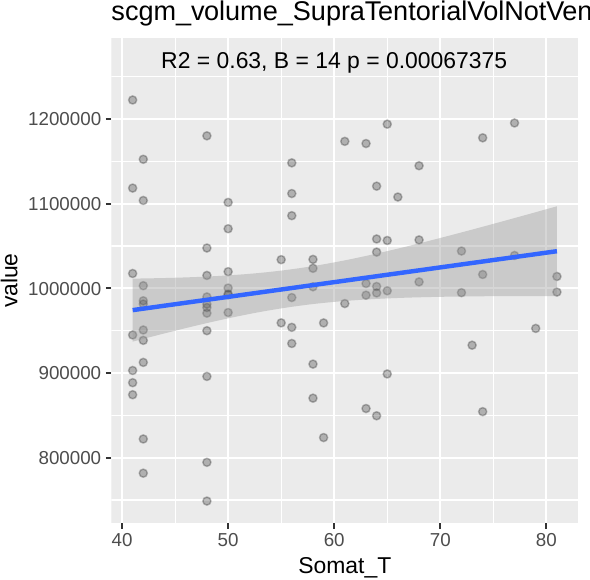

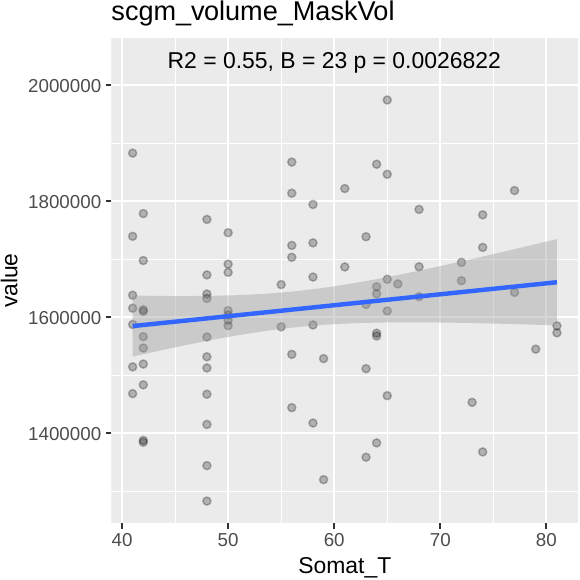

**Figure S4:** Scatterplots indicating the significant relationships between brain areas and somatization scores (here hemispheres are considered separately)

**Table S4**: Life satisfactory uncorrected p-values (hemispheres analyzed together)

**Table S5**: uncorrected p-values of associations between Life Satisfaction and brain regions, analyzing hemispheres separately
